# Temporally annotated textual time series from PubMed Open Access clinical case reports

**DOI:** 10.1101/2025.11.01.25339297

**Authors:** Shahriar Noroozizadeh, Sayantan Kumar, George H. Chen, Jeremy C. Weiss

**Author notes:** These authors contributed equally to this work (Shahriar Noroozizadeh, Sayantan Kumar).

## Abstract

Understanding temporal dynamics in clinical narratives is essential for modeling patient trajectories, yet large-scale temporally annotated resources remain limited. We present PMOA–TTS, the first openly available dataset of 124,699 PubMed Open Access (PMOA) case reports, each converted into structured (event, time) timelines via a scalable LLM-based pipeline. Our approach combines heuristic filtering with Llama 3.3 to identify single-patient case reports, followed by prompt-driven extraction using Llama 3.3 and DeepSeek R1, resulting in over 5.6 million timestamped clinical events. To assess timeline quality, we evaluate against a clinician-curated reference set using three metrics: (i) event-level matching (80% match at a cosine similarity threshold of 0.1), (ii) temporal concordance (c-index > 0.90), and (iii) Area Under the Log-Time CDF (AULTC) for timestamp alignment. Corpus-level analysis shows wide diagnostic and demographic coverage. In a downstream survival prediction task, embeddings from extracted timelines achieve time-dependent concordance indices up to 0.82 *±* 0.01, demonstrating the predictive value of temporally structured narratives. PMOA–TTS provides a scalable foundation for timeline extraction, temporal reasoning, and longitudinal modeling in biomedical NLP. The dataset is available at: https://huggingface.co/datasets/snoroozi/pmoa-tts.

## 1 Introduction

Understanding when clinical events occur is fundamental to modeling patient trajectories, enabling tasks such as process mining, outcome forecasting, and causal inference in medicine [1, 2]. However, most clinical narratives lack explicit temporal structure. While electronic health record (EHR) datasets like MIMIC-III and MIMIC-IV include timestamped structured data and clinical notes [3, 4], they capture only a subset of events (e.g., vital signs, lab results) and still require sophisticated natural language processing to extract temporal information from unstructured text [5, 6].

In contrast, published case reports often provide rich narrative descriptions of patient timelines, but the timing of events is typically expressed in relative terms within the free text (e.g., “on day 3 of hospitalization”). Efforts in temporal reasoning on clinical text have been hampered by the scarcity of large-scale annotated corpora. Earlier initiatives, such as the 2012 i2b2 challenge and Clinical TempEval, focused on temporal relation extraction from clinical notes [7–9] but were constrained by small datasets (e.g., 310 documents) from single institutions, limiting model performance and generalizability. Systems that rely on metadata timestamps (e.g., admission or discharge dates) fail to capture the fine-grained sequence of clinical events described in text, which is essential for nuanced understanding and prediction. There remains a pressing need for large-scale, temporally annotated corpora to support the development and evaluation of models for clinical temporal reasoning.

In this work, we present the PubMed Open Access Textual Time Series (PMOA–TTS) corpus, a new large-scale dataset comprising 124,699 openly available clinical case reports, each annotated with a structured timeline of events. Leveraging large language models (LLMs)—specifically Llama 3.3 [10] and DeepSeek R1 [11]—we transform each case report into a sequence of (clinical event, time) tuples that capture key clinical events and their relative timing within the patient narrative. To our knowledge, PMOA–TTS constitutes the largest publicly available collection of clinical narratives with explicit temporal event structure. We demonstrate that these temporal annotations enable novel forms of analysis and significantly enhance downstream modeling of patient outcomes.

Building PMOA–TTS requires addressing several challenges: identifying relevant case reports at scale, extracting timeline events with high fidelity, and rigorously evaluating the quality of the generated annotations. Rather than relying on naive metadata filters, our LLM-based pipeline accurately identifies single-patient case narratives and resolves ambiguous free-text time expressions to extract fine-grained timelines. We further enrich each case with structured patient demographics and diagnostic labels using prompt-driven extraction. Finally, we showcase the practical value of PMOA–TTS in a downstream survival analysis task, where models leveraging the extracted textual time series demonstrate strong performance in early outcome prediction.

### Contributions

In summary, this paper makes the following contributions:

- We introduce the PMOA Textual Time Series (PMOA–TTS) corpus, comprising 124,699 single-patient case reports from PubMed Open Access, each annotated with structured clinical timelines extracted using large language models (LLMs).
- We develop a scalable LLM-based pipeline for data extraction and annotation, including automatic identification of case reports, prompting strategies for timeline, demographic, and diagnosis extraction, and a manual evaluation framework to assess annotation quality.
- We quantitatively evaluate the accuracy of the extracted timelines, demonstrating high temporal ordering performance (concordance index > 0.9) and introducing a novel Area Under the Log-Time Curve (AULTC) metric for analyzing timing precision and errors.
- We examine demographic distributions, frequently occurring diagnoses, and diagnosis cooccurrence patterns in the corpus, highlighting that it has wide clinical and demographic coverage.
- We show how to use the textual time series in downstream modeling by framing a survival prediction task, where incorporating extracted temporal event sequences improves predictive performance.

PMOA–TTS fills a critical gap by providing a large-scale, openly available resource for temporal reasoning in clinical narratives. We hope this corpus will enable the development and benchmarking of new models for timeline extraction, temporal question answering, and longitudinal outcome prediction in biomedical NLP.

## 2. Related Works

### Clinical Narrative Datasets

Large-scale clinical text data such as those in MIMIC-III [3] and MIMIC-IV [4] have played a pivotal role in clinical NLP research. However, their limitations include a single-institution scope and the absence of temporal event annotations in free-text notes. In contrast, our PMOA–TTS corpus spans a wide range of clinical specialties and institutions globally and provides explicit temporal event sequences extracted from case narratives.

### Biomedical Language Models

Transformer-based models such as BioBERT [12], PubMedBERT [13], and Clinical-T5 [14] have advanced domain-specific understanding of biomedical texts. Still, they are not optimized for temporal reasoning. Our work leverages instruction-tuned LLMs—Llama 3.3 [10] and DeepSeek R1 [11]—to directly generate structured event timelines from unstructured narratives, circumventing the need for intermediate classification steps.

### Temporal NLP Tasks

Temporal information extraction is essential in clinical NLP for modeling patient histories and disease progression. While prior efforts, such as the i2b2 2012 challenge [7], laid foundational work, progress has been constrained by data scarcity. PMOA–TTS supports timeline extraction at scale and introduces novel evaluation metrics like AULTC, bridging the gap between event extraction and downstream temporal modeling (e.g., survival prediction).

### Domain-Specific LLMs

LLMs enable prompt-driven, zero-shot approaches for complex information extraction in healthcare. Domain-adapted models like Med-PaLM [15], GatorTron [16], and BioMedLM [17] extend these capabilities to healthcare applications, with mixed results across health-related tasks. For our pipeline, we apply Llama 3.3 and DeepSeek to tasks including timeline generation and diagnosis identification. A more detailed discussion is presented in Appendix A.

## 3. Methods

Our pipeline comprises the following key components: dataset extraction, textual time series annotation, and comprehensive evaluation of the annotations (Figure 1).

**Figure 1.**
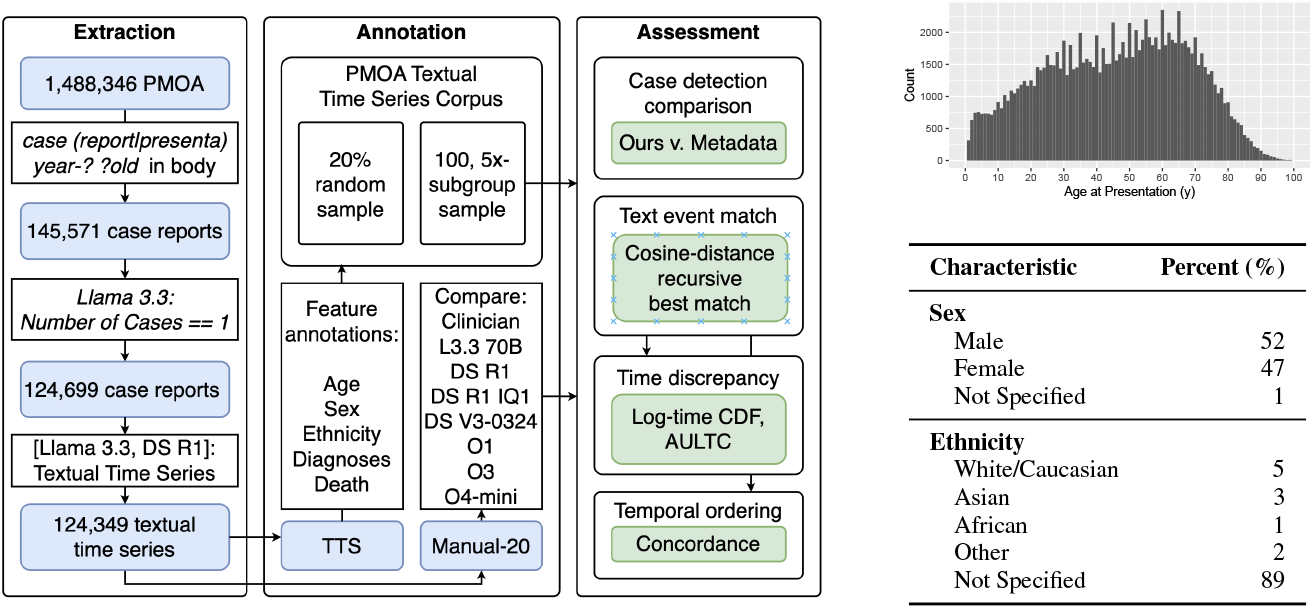
**Left**: Flowchart of the extraction and annotation pipeline. The left panel (Extraction) shows how we filtered the PMOA corpus to identify single-patient case reports. The middle panel (Annotation) depicts the generation of textual time series for each case via LLM prompting and the creation of evaluation subsets. The right panel (Assessment) summarizes the evaluation process, including metadata comparison, text event matching, time discrepancy analysis (log-time CDF, AULTC), and temporal order concordance. **Right**: Age, sex, and ethnicity of the PMOA-TTS corpus.

### 3.1 Case Report Identification Pipeline

We use the PubMed Open Access (PMOA) repository data, which includes 1.4 million published manuscripts. Specifically, we utilize the plain text files from the December 17, 2024 release, located in the `oa_noncomm` folder, which supports both noncommercial and commercial use. Following the method in [18], we identify candidate case reports by extracting content between the section markers “`==== Body`” and “`==== Ref`”, which separate the main text and references in the PMOA format.

To ensure relevance, we first filter documents by matching the body text with the regular expressions “`(case report|case presenta)`” and “`year-? ?old`”. This filter yields 145,571 candidate documents likely to be case reports or case series. To narrow this set to single-patient case reports, we use a large language model (LLaMA 3.3 70B Instruct) to analyze each document’s body text. The model is prompted with: “`You are a physician. Determine the number of case reports in the following manuscript. Return 0 if it is not a case report. Reply only with the number and nothing else.`”, followed by the document body. We retain documents for which the model replies with 1 and discard others. This two-stage process results in a final dataset of 124,699 single-patient case reports (Figure 1).

We compared our LLM-based case report identification method with PubMed’s metadata-based labeling using a carefully designed benchmark. We created a dataset of 100 reports across five diagnoses—Diabetes, Sepsis, Hypertension, COVID-19, and Atrial Fibrillation—with 10 reports per diagnosis from each method. A clinician manually reviewed and labeled each report to verify whether it described exactly one case. Our LLM-pipeline significantly outperforms PubMed metadata filtering, particularly in terms of specificity and precision (Section 4.2).

### 3.2 Timeline Annotation via LLM Prompting

We used LLMs to extract clinical textual time series from PMOA case reports. A textual time series is a chronologically ordered sequence of clinical findings, each associated with a timestamp and a specific patient. A clinical finding refers to a free-text description of a health-related condition affecting the patient. Our approach differs from conventional methods (e.g., [7, 19]) by capturing longer, more context-rich text spans to accurately convey the meaning of clinical findings. Unlike the i2b2 guidelines, we do not limit findings to short prepositional phrases. For example, “pain in chest that radiates substernally” is kept intact to preserve its clinical nuance. Additionally, coordinated findings are separated for clarity—e.g., “metastases in the liver and pancreas” is split into “metastasis in the liver” and “metastasis in the pancreas.”

To construct clinical textual time series, we adopt the LLM extraction prompt from previous work [18, 20]. This prompt instructs the LLM to extract clinical events with timestamps, setting the admission event at a reference time of 0, with other events expressed in relative hours (negative for pre-admission events, positive for post-admission). It also provides guidelines for handling conjunctions, event durations, and output formatting (full prompt in Appendix B.1).

For each report, the LLM generates a structured sequence of (clinical_event, time) tuples. Events are retained in free-text form to preserve their clinical specificity, rather than being mapped to a fixed ontology. On average, each report contains 46 events, with significant variation depending on report length and complexity. The corpus as a whole includes over 5.6 million temporally anchored events, making it one of the largest datasets of its kind in clinical NLP. Computational resources used are described in Appendix J.

### 3.3 Manual Annotations for Evaluation

We combined manual annotations from [18] and [20] to serve as ground truth for evaluating the quality of LLM annotations. The manual annotation process was conducted by expert physicians and followed the ISO-TimeML specification, which includes actions or states inferred to have occurred or held true. Annotators selected exact text spans representing events, with two exceptions to enhance usability: (i) conjunctive lists could be split into individual events, and (ii) the prefix “history of” could be added when contextually appropriate. These adjustments preserved the original text alignment for downstream analysis. Each event was then assigned a time relative to the case reference point (e.g., admission or encounter date), using the start time when an interval was described.

This manually annotated subset of 20 cases serves as the gold standard for our evaluation metrics, described in the following section.

### 3.4 Evaluation Metrics

We assessed the textual time series extracted from PMOA case reports using three key metrics: event match rate, temporal concordance (c-index), and log-time discrepancy.

#### Text Event Match

To evaluate the alignment between predicted clinical findings and reference annotations, we used a recursive best-match strategy [18, 20]. This method pairs events by minimizing string-based distances while avoiding many-to-one mappings. For each reference event, the closest predicted event is selected based on the minimal distance, with event order used to break ties. Once matched, both events are removed from further consideration, and the process continues recursively with the remaining pairs.

We tested several distance metrics, including Levenshtein distance, cosine similarity of BERT (bertbase-uncased) embeddings, and cosine similarity based on sentence embeddings from PubMedBERT (S-PubMedBert-MS-MARCO). Previous validation showed that PubMedBERT-based cosine similarity with a threshold of 0.1 was the most reliable, and we used this metric for our evaluation. A detailed description of the algorithm is provided in Appendix E.

#### Temporal Ordering

To quantify the accuracy of temporal ordering, we used the concordance index (c-index), which measures the probability that a randomly selected pair of predicted findings is correctly ordered in time. Higher c-index values indicate better alignment with the reference timeline. Formally, for a set of events with reference times 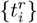 and predicted time 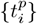, the c-index is calculated as 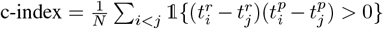 where *N* is the number of comparable pairs (event indices *i, j* such that 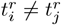 and 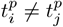), and 𝟙{·} is the indicator function.

#### Time Discrepancy Analysis

We evaluated temporal prediction accuracy by measuring the discrepancies between predicted and reference timestamps. Log-time discrepancy was computed as log(1 + Δ*t*), where Δ*t* is the absolute time difference (in hours). To summarize these discrepancies, we introduced the Area Under the Log-Time Cumulative Distribution Function (AULTC), which quantifies how quickly the cumulative distribution of time errors rises—higher AULTC values indicate better temporal alignment. The full definition of AULTC is in Appendix D. We also stratified the analysis by reference time intervals (hour, day, week, year) to examine how prediction accuracy varies with time from case presentation.

### 3.5 Downstream Applications

To demonstrate the utility of the PMOA-TTS corpus for clinical prediction tasks, we implemented two downstream applications: survival analysis and event forecasting.

#### Survival Analysis

An example downstream task enabled by PMOA-TTS is survival analysis, where the objective is to predict time until death extracted from each case report. We adopt a twostage framework: first, each textual timeline is converted into a structured context of timestamped clinical events (following the Triplet encoding approach in [21]) and embedded using various pretrained encoder and decoder language models. For encoder models, we extract the [CLS] token embedding; for decoder models, we compute the mean over non-padding token embeddings. Second, these representations serve as covariates for two survival models: DeepSurv [22] and DeepHit [23]. Performance is evaluated using the time-dependent concordance index [24] and the integrated Brier score, with hyperparameter tuning on a validation set and final evaluation on a held-out test set. To ensure robustness, experiments were repeated across five random seeds. Full experimental details are provided in Appendix H.

#### Event Forecasting Applications

In parallel work by [25], the utility of textual time series for forecasting tasks has been demonstrated on a smaller cohort of sepsis patients. That work formulated two specific forecasting tasks:

1. *Next Event Timing (F1)*: Given a history of (clinical_event, time) tuples, predicting whether the next *k* events will occur within specific time windows (1 hour, 1 day, 1 week).
2. *Event Ordering (Concordance)*: Given a history of (clinical_event, time) tuples, predicting the correct temporal ordering of the next *k* events.

A summary of these forecasting tasks and their experimental results from the PMOA-TTS subset used in [25] are provided in Appendix I for completeness.

## 4 Results

### 4.1 Demographic Characterization of PMOA-TTS

For demographic extraction, we used an LLM-based query to retrieve age, sex, and ethnicity to characterize the demographic composition of the corpus. The prompt specified that age be reported in years, sex as Male, Female, a custom string, or Not Specified, and ethnicity according to U.S. Census categories or as Not Specified (see Appendix B.2).

Our analysis shows a median age of 47 years, with age distribution peaks at 5-year intervals, indicating rounding in clinical reporting (Figure 1). The gender distribution is balanced (52% male, 47% female, and 1% with unspecified gender.) Most case reports (89%) do not specify the patient’s ethnicity. When ethnicity is specified, White/Caucasian (5%) and Asian (3%) are the most common, followed by African (1%) and Other (2%). Compared to U.S. population demographics, Asian patients are overrepresented, likely due to the international scope of PubMed publications.

### 4.2 Case Report Identification Performance

Our evaluation of case report identification methods compared our LLM-Pipeline approach against PubMed’s metadata-based filtering across five different diagnoses: Diabetes, Sepsis, Hypertension, COVID-19, and Atrial Fibrillation. Using clinician-assigned labels as ground truth, we computed precision, recall, accuracy, and F1 score for each approach.

The LLM-Pipeline approach demonstrates superior performance with perfect precision (1.00) across all diagnostic categories and also aggregated across all diagnoses (Table 1), indicating that when our pipeline identifies a document as a single-patient case report, it is always correct. In contrast, the PubMed MetaData approach shows more variable precision, ranging from 0.84 for Atrial Fibrillation to 1.00 for Hypertension, with an overall precision of 0.92.

**Table 1:** Case report identification performance per diagnosis and aggregated across all diagnoses.

| Subset | Diagnosis | Precision | Recall | Accuracy | F1 Score |
| --- | --- | --- | --- | --- | --- |
| PubMed MetaData | Atrial Fibrillation | 0.84 | 0.94 | 0.80 | 0.89 |
|  | COVID-19 | 0.94 | 0.89 | 0.85 | 0.92 |
|  | Diabetes | 0.85 | 1.00 | 0.85 | 0.92 |
|  | Hypertension | 1.00 | 1.00 | 1.00 | 1.00 |
|  | Sepsis | 0.95 | 0.95 | 0.90 | 0.95 |
|  | **All Diagnoses** | 0.92 | **0.96** | 0.88 | 0.94 |
| LLM-Pipeline | Atrial Fibrillation | 1.00 | 1.00 | 1.00 | 1.00 |
|  | COVID-19 | 1.00 | 0.89 | 0.90 | 0.94 |
|  | Diabetes | 1.00 | 0.94 | 0.95 | 0.97 |
|  | Hypertension | 1.00 | 0.95 | 0.95 | 0.97 |
|  | Sepsis | 1.00 | 1.00 | 1.00 | 1.00 |
|  | **All Diagnoses** | **1.00** | **0.96** | **0.96** | **0.98** |

Both methods achieve similar recall (0.96), indicating comparable success in identifying true case reports. However, the LLM-Pipeline attains higher overall accuracy (0.96 vs. 0.88), reflecting its superior ability to distinguish both true positives and true negatives. This accuracy gain is largely due to the LLM-Pipeline’s effectiveness in excluding non-single-patient reports, which the PubMed MetaData approach often misclassifies.

Examining performance by diagnostic category reveals that both approaches perform best on Sepsis and Hypertension cases, while showing more variability with Atrial Fibrillation, COVID-19, and Diabetes reports. The LLM-Pipeline maintains consistently high performance across all categories, with F1 scores ranging from 0.94 to 1.00. These findings highlight the significant improvements in precision and overall classification quality offered by our LLM-Pipeline approach for case report identification, ensuring that the PMOA-TTS corpus contains high-quality, single-patient case reports with minimal contamination from multi-patient or non-case report documents.

### 4.3 Evaluation of Clinical Textual Time Series Quality

We evaluated the quality of extracted textual time series using the manually annotated subset as ground truth, comparing seven LLM variants: DeepSeek-R1, DeepSeek-R1-UD-IQ1, DeepSeek-V3-0324, LLaMA 3.3 70B, OpenAI O1, OpenAI O3, and OpenAI O4-mini. A comparison of different LLM annotations and sensitivity to different prompt strategies can be found in Appendices C and K respectively.

#### Text Event Match Performance

Figure 2 (left) shows the cumulative density of cosine distances between model-generated and reference event descriptions. A threshold of 0.1 (vertical line) defines a successful match. DeepSeek-R1 (black) and its quantized variant DeepSeek-R1-UD-IQ1 (orange) outperform other models, with 80% of aligned events falling below the threshold. DeepSeek-V3-0324 (light blue) and O4-mini (red) form a second tier, while LLaMA 3.3 70B (green), O1 (light yellow), and O3 (blue) show more dispersed distributions, with only 70% of events below the threshold. These analyses highlights clear differences in how well models generate event descriptions that align semantically with expert annotations. The strong performance of DeepSeek-R1 and its quantized variant indicates their enhanced ability to model clinical language patterns found in case reports.

**Figure 2.**
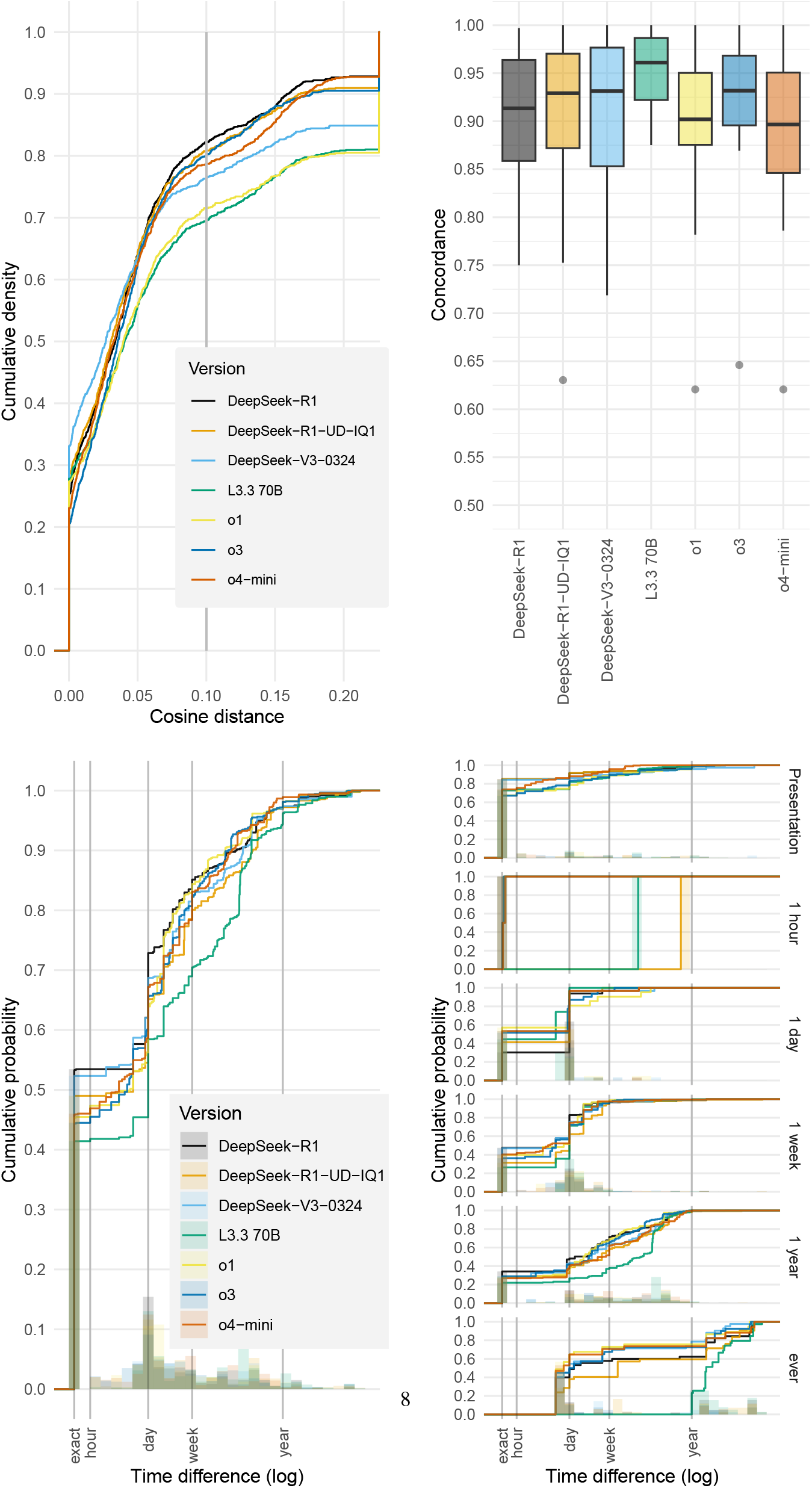

#### Temporal Ordering Performance

Using the cosine distance threshold to identify matched events, we assessed temporal ordering accuracy via the concordance index (c-index). As shown in Figure 2 (right), LLaMA 3.3 70B achieves the highest median c-index (0.96), with DeepSeek-V3-0324 and DeepSeek-R1 following closely.

The relationship between event matching and concordance evaluation is important to consider. Models applying stricter matching thresholds may appear to perform better on concordance by evaluating only the most confidently matched events. In contrast, models matching a broader set of events—including more ambiguous cases—encounter a more difficult ordering task. LLaMA 3.3 70B’s strong performance in temporal ordering is notable given its lower match rate, suggesting it effectively captures relative timing even when its textual matches are less precise. Appendix F discusses how adjusting the event match threshold allows tuning the balance between match coverage and ordering accuracy.

#### Time Discrepancy Analysis

Figure 2 (left) displays the cumulative probability distribution of time differences (log-scaled) between predicted and reference timestamps, illustrating each model’s timestamping accuracy. DeepSeek-R1 (black line) shows the steepest curve, indicating that a higher proportion of its predictions are temporally close to the reference points. In contrast, LLaMA 3.3 70B (green line) produces a flatter curve, reflecting greater variation and lower precision in absolute time prediction. The right panel in Figure 2 breaks down performance across time intervals—presentation, 1 hour, 1 day, 1 week, 1 year, and beyond. All models perform well for events near the presentation time, but their accuracy declines as the temporal distance increases. DeepSeek models consistently show stronger performance across intervals, whereas LLaMA 3.3 70B exhibits more noticeable degradation for distant events.

Taken together, this analysis highlights a key distinction: while LLaMA 3.3 70B is more effective at predicting relative event order (concordance), DeepSeek models—particularly DeepSeek-R1—are better at assigning precise timestamps, even for events far from the reference point.

### 4.4 Diagnosis Distribution Analysis

Additionally, we prompted the LLM to act as an expert physician and list patient-specific diagnoses from the report, distinguishing true diagnoses (e.g., “acne,” “DRESS syndrome”) from general clinical findings (e.g., “rash,” “leukocytosis”). The prompt instructed the model to list the primary diagnosis first, include one diagnosis per line, and omit explanatory text (full prompt in Appendix B.3).

To assess the medical coverage of our corpus, we analyzed the diagnoses extracted through this LLM-based method, as detailed in Appendix G. Figure 3a shows the 20 most frequent diagnoses, demonstrating the broad clinical scope of the dataset. Hypertensive disorders, including hypertensive disease, diastolic and systolic hypertension, prehypertension, and renal hypertension, are the most common, followed by diabetes mellitus and its subtypes.

**Figure 3.**
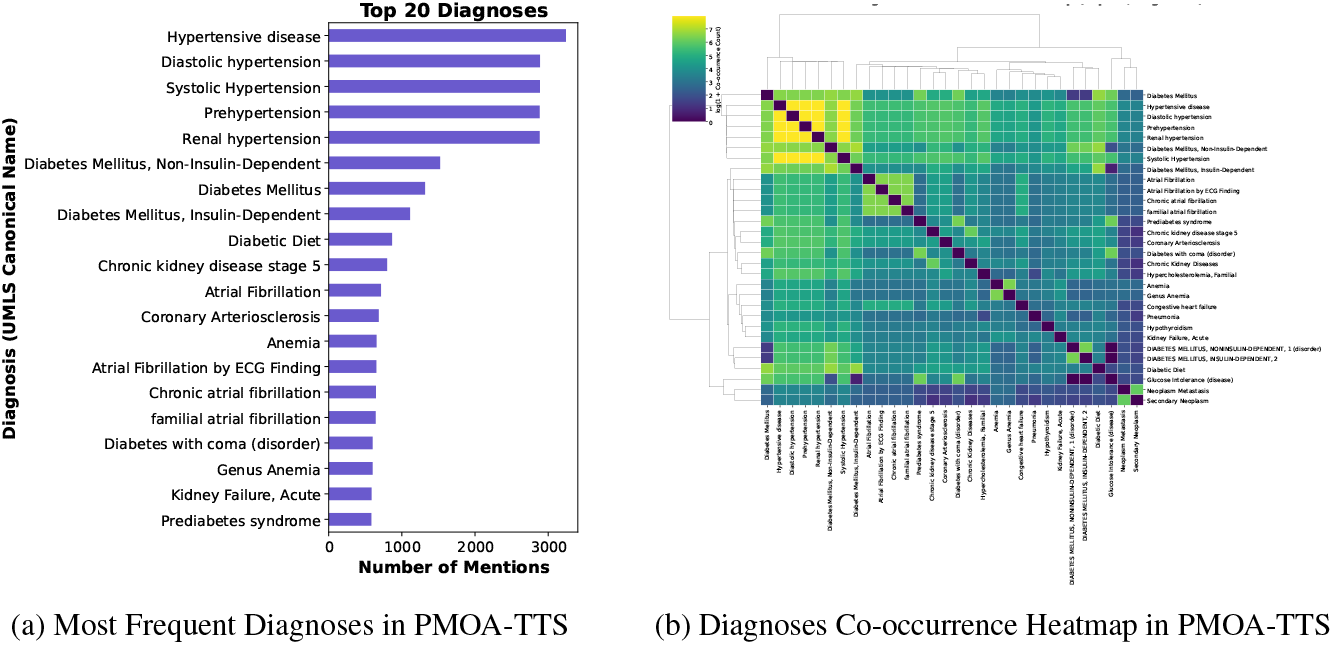
Co-occurrence Patterns and Frequency of Diagnoses in Clinical Case Reports. The left panel shows the top 20 most frequently mentioned diagnoses, using UMLS canonical names, across clinical case reports. Cardiovascular and metabolic conditions such as hypertension, diabetes mellitus, and chronic kidney disease dominate the list. The right panel is a heatmap with hierarchical clustering, visualizing pairwise co-occurrence patterns (log-transformed counts) among diagnoses. Off-diagonal values reveal common comorbidity patterns in our PMOA-TTS. Clustering reveals distinct groups of frequently co-mentioned conditions, such as cardiovascular disorders and diabetes-related terms.

This distribution aligns with the prevalence of chronic conditions typically seen in clinical practice, particularly cardiovascular, metabolic, and renal disorders, and matches the corpus’s demographic profile, with a median patient age of 45 years—an age group commonly affected by these conditions.

We also examined diagnosis co-occurrence patterns using a clustered heatmap (Figure 3b), revealing strong groupings of related conditions. Notably, hypertensive spectrum disorders and diabetes variants cluster together, reflecting well-established clinical links, such as those between diabetes and chronic kidney disease, or atrial fibrillation and hypertension.

This analysis highlights the diverse range of medical conditions in the PMOA-TTS corpus and demonstrates its suitability for clinical prediction tasks across multiple disease domains.

### 4.5 Survival Analysis Results

Table 2 presents the performance of different language model embeddings and survival models in predicting patient outcomes across three time thresholds (0h, 24h, and 168h). We show results for the time-dependent concordance index, which measures the model’s ability to correctly rank patients by their risk. Descriptive analysis is provided in Appendix H.

**Table 2:** Time-dependent concordance (mean ± standard deviation across 5 experimental runs) index across embedding models and survival modeling approaches at different time thresholds.

| Embedding Model | Time Threshold = 0h |  | Time Threshold = 24h |  | Time Threshold = 168h |  |
| --- | --- | --- | --- | --- | --- | --- |
|  | DeepHit | DeepSurv | DeepHit | DeepSurv | DeepHit | DeepSurv |
| DeepSeek-R1-Distill-Llama-70B | 0.815 ± 0.010 | 0.812 ± 0.013 | 0.815 ± 0.012 | 0.818 ± 0.009 | 0.810 ± 0.012 | 0.812 ± 0.014 |
| DeepSeek-R1-Distill-Llama-8B | 0.806 ± 0.015 | 0.803 ± 0.011 | 0.809 ± 0.013 | 0.806 ± 0.011 | 0.802 ± 0.010 | 0.809 ± 0.011 |
| Llama-3.1-8B-Instruct | 0.815 ± 0.008 | 0.813 ± 0.010 | 0.817 ± 0.010 | 0.811 ± 0.007 | 0.808 ± 0.014 | 0.809 ± 0.011 |
| Llama-3.3-70B-Instruct | **0.822 ± 0.004** | **0.817 ± 0.011** | **0.821 ± 0.011** | **0.819 ± 0.005** | **0.819 ± 0.009** | **0.822 ± 0.009** |
| ModernBERT-base | 0.718 ± 0.011 | 0.729 ± 0.008 | 0.733 ± 0.010 | 0.738 ± 0.011 | 0.725 ± 0.013 | 0.729 ± 0.009 |
| ModernBERT-large | 0.653 ± 0.014 | 0.667 ± 0.009 | 0.655 ± 0.015 | 0.665 ± 0.017 | 0.643 ± 0.011 | 0.662 ± 0.003 |
| bert-base-uncased | 0.743 ± 0.013 | 0.746 ± 0.009 | 0.750 ± 0.014 | 0.748 ± 0.006 | 0.744 ± 0.009 | 0.747 ± 0.010 |
| deberta-v3-small | 0.739 ± 0.010 | 0.753 ± 0.010 | 0.730 ± 0.019 | 0.758 ± 0.006 | 0.733 ± 0.012 | 0.751 ± 0.006 |
| roberta-base | 0.757 ± 0.017 | 0.756 ± 0.013 | 0.760 ± 0.012 | 0.758 ± 0.014 | 0.761 ± 0.009 | 0.762 ± 0.011 |

Several important patterns emerge from our results. Decoder-based models (DeepSeek, LLaMA) consistently outperform encoder-based models (BERT variants) across all time horizons and survival modeling approaches. LLaMA-3.3-70B-Instruct delivers the strongest performance overall, achieving concordance indices above 0.82 in most scenarios. Within each model family, larger architectures (e.g., 70B vs. 8B) generally yield superior results, suggesting that greater model capacity enhances embedding quality for downstream survival prediction. Regarding observation windows, predictions that incorporate longer time spans (24h, 168h) show modest gains over the baseline (0h), indicating that additional temporal context contributes to improved prognostic accuracy, albeit with a relatively small effect. In terms of survival modeling frameworks, DeepSurv and DeepHit exhibit comparable performance across most conditions, with no consistent advantage for either method.

Overall, these findings indicate that high-quality embeddings from LLMs can effectively encode prognostic signals from textual time series, with decoder-based models in particular showing strong potential for survival prediction tasks.

## 5 Discussion and Conclusion

This work introduces PMOA–TTS, a large-scale corpus of 124,699 temporally annotated clinical case reports generated using large language models (LLMs). Our method transforms unstructured narratives into structured temporal sequences, addressing a key challenge in biomedical NLP. Evaluation against expert annotations shows that LLM-based extraction achieves high accuracy in event detection and temporal ordering, enabling scalable clinical timeline analysis. Our LLM-based pipeline for identifying case reports outperforms traditional metadata filtering, marking a methodological advancement with wider applicability. With near-perfect precision (1.00) and high recall (0.96), it reliably selects genuine single-patient case reports—crucial for preserving the integrity of downstream temporal analyses. These results indicate that LLM-based filtering can improve corpus quality across other medical document types where metadata alone falls short.

Comparative analysis of LLMs reveals key differences in temporal reasoning. DeepSeek models—especially DeepSeek-R1—show stronger semantic alignment with expert annotations in event matching. Our results highlight complex interactions between concordance and time discrepancy metrics, with the event matching threshold significantly influencing which events are included in concordance evaluations. Stratified analysis also shows that all models perform worse on events further from the reference point, indicating a crucial area for future improvement.

Our diagnosis mapping and demographic profiling support the creation of curated cohorts from the full corpus. Researchers can easily construct disease-specific (e.g., diabetes, hypertension, atrial fibrillation) or demographic-based subgroups for targeted temporal studies without needing new data. For example, they can analyze progression in rare diseases or compare timelines across different populations. With over 5.6 million temporally annotated events, the corpus enables statistically robust analyses across a wide range of clinical conditions.

Downstream survival modeling shows that LLMs vary in how effectively they encode temporal information from clinical narratives. Decoder-based models (LLaMA, DeepSeek) consistently outperform encoder-based ones (BERT variants) across time horizons and modeling methods. LLaMA-3.3-70B-Instruct achieves the highest concordance, indicating that larger models better capture temporal complexity. While the differences between survival frameworks (DeepSurv vs. DeepHit) are minor, the language model choice significantly impacts prediction accuracy. This analysis showcases just one application of PMOA–TTS, underscoring its value for diverse temporal prediction tasks. Our reference to concurrent work [25] on temporal forecasting underscores both the potential and limitations of clinical narrative prediction. The underperformance of general-purpose LLMs highlights the need for models tailored to temporal reasoning. PMOA–TTS provides a robust testbed for advancing such models, spanning a broad spectrum of clinical conditions beyond domains like sepsis.

### Limitations and Future Directions

This study has several key limitations. First, we model events as minimally modified text spans to preserve alignment with the source text, which facilitates interpretability but relies heavily on embedding-based comparison and forgoes normalization to clinical ontologies. Second, representing time with single relative points simplifies annotation but limits expressiveness, especially in cases where durations or implicit temporal relationships are critical. Third, our one-to-one recursive event matching strategy prioritizes temporal consistency, but at the cost of reduced recall, particularly when events are expressed as conjunctive or split mentions. These design choices emphasize precision and contextual grounding but leave room for enhancements in coverage and temporal fidelity. Future work could incorporate LLM ensembling, prompt engineering, or active learning-based fine-tuning to further improve event-time extraction. Ultimately, we envision the PMOA–TTS corpus as a valuable foundation for downstream applications such as forecasting, process discovery, and AI-augmented clinical reasoning. By introducing a large, publicly available resource with detailed temporal annotations, this work enables scalable investigations of disease progression and time-based phenotypes, with the potential to generalize to other clinical documents and specialized temporal tasks. A more detailed discussion about the positive and negative societal impacts of our work have been presented in Appendix L.

## Data Availability

The dataset is derived from publicly available PubMed Open Access case reports. All annotations and metadata are released under the CC BY NC SA 4.0 license.

https://huggingface.co/datasets/snoroozi/pmoa-tts

https://github.com/jcweiss2/pmoa_tts

## Acknowledgments

This research was supported in part by the Division of Intramural Research (DIR) of the National Library of Medicine (NLM), National Institutes of Health. This work utilized the computational resources of the NIH HPC Biowulf cluster.

## Appendix

This Appendix provides supplementary material supporting the main text, including implementation details, additional evaluations, and methodological clarifications. It is intended to enhance reproducibility and transparency by including full prompt templates, evaluation procedures, metric definitions, model settings, and examples. It also extends the main findings with additional analyses such as prompt ablation studies, threshold sensitivity, UMLS normalization, and forecasting on condition-specific subsets. The Appendix is organized into thematic sections, each described below.

### Appendix A: Related Works

This section briefly reviews prior work in clinical timeline extraction, including benchmarks like i2b2 and THYME. It situates PMOA–TTS within this context and highlights the lack of large-scale open-access corpora for temporal clinical NLP.

### Appendix B: Prompt Templates for Annotation Tasks

This section includes the full prompt used for timeline extraction, including detailed instructions and few-shot examples. It also presents the LLM prompts used for extracting demographics and diagnoses from case reports, which are essential for downstream characterization.

### Appendix C: Comparing LLM Annotations

Here, we show example outputs from different LLMs alongside manual annotations to highlight differences in event span selection and temporal alignment. These side-by-side examples help illustrate the semantic variation across models.

### Appendix D: Log-Time Cumulative Distribution Function

This section introduces the AULTC (Area Under the Log-Time CDF) metric for evaluating absolute timestamping error. It explains how this metric captures the cumulative deviation between predicted and reference times on a log-transformed scale.

### Appendix E: Recursive Best Match Procedure

We describe the recursive algorithm used to align predicted and reference events during evaluation. The method ensures one-to-one matching using semantic similarity scores and resolves ties based on event order.

### Appendix F: Varying the Event Match Distance Threshold

This section explores the sensitivity of evaluation metrics (match rate, concordance, AULTC) to different cosine similarity thresholds for event matching. The analysis helps quantify trade-offs between match precision and coverage.

### Appendix G: UMLS Mapping for Diagnosis Standardization

We describe the process of linking free-text diagnoses to standardized UMLS concepts using ScispaCy. This mapping enables systematic analysis of disease frequency and co-occurrence patterns across the corpus.

### Appendix H: Survival Analysis Task on PMOA-TTS

This section details the setup and hyperparameters for the survival prediction task using textual time series embeddings. It includes information on model architectures, embedding strategies, training regimes, and evaluation metrics.

### Appendix I: Forecasting Task on the Sepsis Subset of PMOA-TTS

We reproduce results from a prior study using a subset of sepsis case reports to demonstrate forecasting of future clinical events. These findings serve as a proof-of-concept for temporal prediction using PMOA–TTS.

### Appendix J: Computational Resources for Reproducibility

This section outlines the hardware and compute environment used for LLM inference, survival modeling, and evaluation. It includes GPU types, inference times, and total compute hours for each major pipeline stage.

### Appendix K: Sensitivity to the Quality of LLM Prompts

We evaluate how the inclusion or removal of specific prompt components (e.g., role definition, examples, conjunction splitting) affects model performance. The section includes quantitative comparisons across prompt variants.

### Appendix L: Broader Impact

This section discusses both the potential benefits and risks associated with the release of PMOA–TTS. Topics include improved accessibility and modeling capabilities, as well as concerns about bias, data quality, and misuse in clinical contexts.

### A. Related Works

#### Clinical Narrative Datasets

Large corpora of clinical text have enabled significant advances in biomedical NLP. The MIMIC-III [3] and MIMIC-IV [4] databases of de-identified hospital ICU records are among the most widely used resources, containing clinical notes and structured data for hundreds of thousands of patient stays. These datasets include timestamped events such as lab tests and interventions, which have been used for sequential modeling and outcome prediction [26, 27]. However, they are limited to a single institutional setting, and the free-text notes in MIMIC are not annotated for temporal event structures. In contrast, our PMOA-TTS corpus draws from published case reports across diverse institutions and specialties worldwide, providing a broad distribution of patient scenarios. Additionally, prior to our work, no large open-access corpus existed with extracted temporal sequences from narrative texts. Smaller annotated corpora for clinical temporal reasoning have been developed, such as the i2b2 2012 Temporal Relations challenge corpus [7], the THYME corpus of oncology notes [28], and MedTimeML [29], but these contain only a few hundred documents and require special access. PMOA-TTS dramatically scales up the availability of temporally annotated clinical narratives (by two orders of magnitude in document count) and is freely usable, facilitating LLM approaches.

#### Biomedical Language Models

The success of transformer language models pretrained on large corpora has encouraged the creation of domain-specific models for biomedicine. BioBERT [12], PubMedBERT [13], and BioELECTRA [30] are encoder-only transformer models trained on PubMed abstracts and full-text articles, achieving strong results on biomedical NLP benchmarks by capturing domain-specific terminology and semantics. [31] further incorporates clinical notes (MIMIC-III) into pretraining, bridging biomedical literature and clinical text. Recent advancements include BioGPT [32] and Clinical-T5 [14], which extend the decoder-only and encoder-decoder paradigms to biomedical text understanding and generation. These models provide powerful text representations for static biomedical text understanding. However, modeling temporal information in narratives poses additional challenges, as it requires capturing event order and timing, not just context at the sentence or document level [33, 34]. Some recent works attempt to adapt language models for clinical timeline tasks. For example, approaches for temporal relation extraction in clinical text have combined rule-based and learned methods [18], and sequence-to-sequence and classification strategies have been applied to link events with time expressions [35–38]. Our work differs in that we leverage large instruction-tuned models—specifically Llama 3.3 [10] and DeepSeek R1 [11]—to directly generate structured timeline annotations from text, rather than classifying links. We also introduce an end-to-end pipeline that can process hundreds of thousands of documents, demonstrating the scalability of LLMs for corpus-wide annotation.

#### Temporal NLP Tasks

Beyond the clinical domain, temporal information extraction has been explored in newswire and narrative text (e.g., the TimeBank corpus [39], TempEval challenges [40], and temporal evaluation benchmarks [41]). These typically involve identifying time expressions, events, and temporal relations (before/after) to construct timelines. In the clinical domain, understanding patient timelines has clear importance for tasks like disease progression modeling and treatment effect monitoring [42, 43]. The 2012 i2b2 challenge [7] established baseline techniques for extracting events, temporal expressions, and relations from discharge summaries, which have been extended by subsequent works on clinical timeline visualization and timeline reconstruction [38, 44, 45]. While those efforts advanced the field, progress has been limited by data availability. Our PMOA-TTS dataset contributes to temporal NLP by providing a large-scale testbed specifically focused on clinical case narratives. It also enables new problem formulations; for instance, we treat timeline extraction as a structured generation task using LLMs, and we evaluate temporal extraction quality with novel metrics like Area Under the Log-Time Curve (AULTC). Moreover, we show how the extracted timelines can feed into downstream temporal modeling (survival analysis), connecting information extraction with predictive analytics. This bridges the gap between temporal NLP and clinical outcome modeling, an intersection that has been relatively under-explored.

#### Domain-Specific LLMs

The recent emergence of large language models (LLMs) has opened avenues for zero-shot and few-shot information extraction without task-specific training. Models such as GPT-3/4 [46, 47], LLaMA [48], and DeepSeek [11] demonstrate the ability to follow prompts and generate structured outputs, including in the medical domain. Domain-adapted models like Med-PaLM [15], GatorTron [16], and BioMedLM [17] further extend these capabilities specifically for healthcare applications. Researchers have begun to apply LLMs to tasks like clinical concept extraction [49], medical question answering [15], and discharge summary generation [50] with promising results. In this work, we leverage LLMs—specifically Llama 3.3 [10] and DeepSeek [11]—to perform complex annotation tasks (identifying case reports, extracting timeline events, and also downstream identifying diagnosis for each case report) by simply providing carefully designed prompts. We compare these LLM variants in our pipeline for their efficacy in timeline extraction, contributing to understanding the capabilities and limitations of current LLMs on temporal reasoning tasks. Additionally, we highlight the need for high-quality LLM annotations: since these models are not perfect, their output can introduce noise that impacts downstream models [51]. By quantifying LLM annotation accuracy and its effect on a survival prediction task, we underscore the importance of improving LLM-based extraction (through better prompts or model improvements) for high-stakes clinical applications.

### B Prompt Templates for Annotation Tasks

#### B.1 LLM prompt to generate textual time-series from PMOA case reports

##### Prompt

You are a physician. Extract the clinical events and the related time stamp from the case report. The admission event has timestamp 0. If the event is not available, we treat the event, e.g. current main clinical diagnosis or treatment with timestamp 0. The events that happened before event with 0 timestamp have negative time, the ones after the event with 0 timestamp have positive time. The timestamp are in hours. The unit will be omitted when output the result. If there is no temporal information of the event, please use your knowledge and events with temporal expression before and after the events to provide an approximation. We want to predict the future events given the events happened in history. For example, here is the case report.

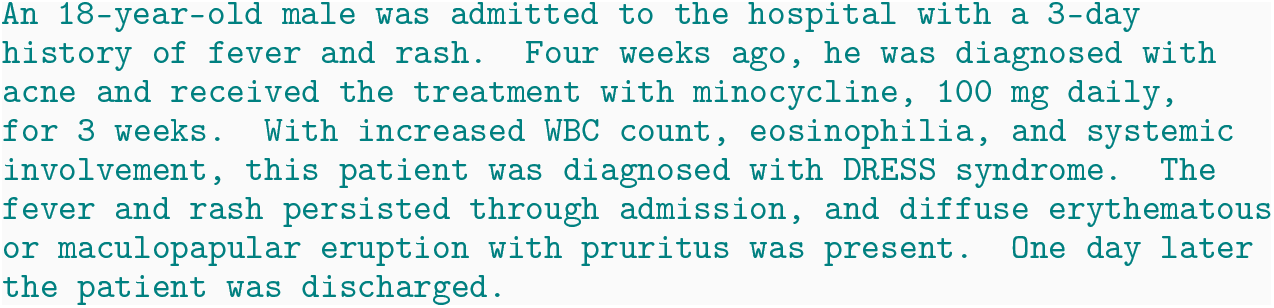

Let’s find the locations of event in the case report, it shows that four weeks ago of fever and rash, four weeks ago, he was diagnosed with acne and receive treatment. So the event of fever and rash happened four weeks ago, 672 hours, it is before admitted to the hospital, so the time stamp is −672. diffuse erythematous or maculopapular eruption with pruritus was documented on the admission exam, so the timestamp is 0 hours, since it happens right at admission. DRESS syndrome has no specific time, but it should happen soon after admission to the hospital, so we use our clinical judgment to give the diagnosis of DRESS syndrome the timestamp 0. then the output should look like:

~~~
18 years old | 0
male | 0
admitted to the hospital | 0 fever | −72
rash | −72
acne | −672
minocycline | −672
increased WBC count | 0
eosinophilia| 0
systemic involvement| 0
diffuse erythematous or maculopapular eruption| 0
pruritis | 0
DRESS syndrome | 0
fever persisted | 0
rash persisted | 0
discharged | 24
~~~

Separate conjunctive phrases into its component events and assign them the same timestamp (for example, separation of ‘fever and rash’ into 2 events: ‘fever’ and ‘rash’). If the event has duration, assign the event time as the start of the time interval. Attempt to use the text span without modifications except ‘history of’ where applicable. Include all patient events, even if they appear in the discussion; do not omit any events; include termination/discontinuation events; include the pertinent negative findings, like ‘no shortness of breath’ and ‘denies chest pain’. Show the events and timestamps in rows, each row has two columns: one column for the event, the other column for the timestamp. The time is a numeric value in hour unit. The two columns are separated by a pipe ‘|’ as a bar-separated file. Reply with the table only.

#### B.2 LLM prompt to extract demographics from PMOA case reports

##### Prompt

You are an expert clinician. Extract the following demographic information from the case report: (1) age at case presentation, (2) sex (biologic sex at birth), and (3) ethnicity. Report the age in years. Report the sex as Male, Female, otherwise as a custom string, or Not Specified. Report ethnicity as according to the US Census, otherwise as Not Specified. For example, here is the case report.

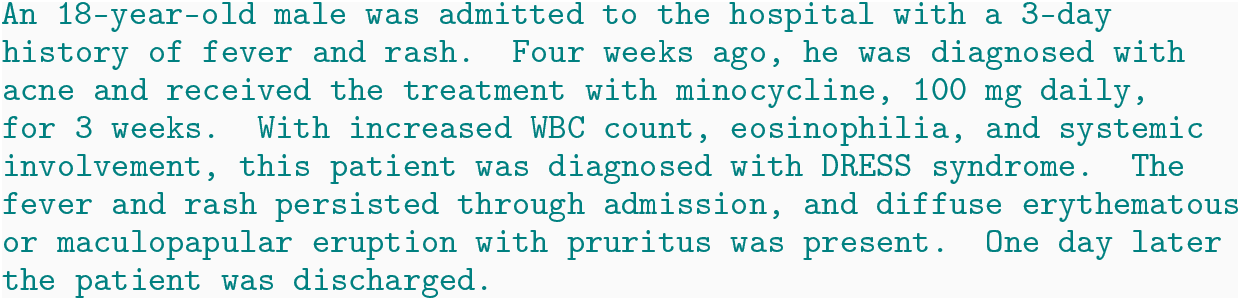

Then the demographic extraction should be:

~~~
18 | Male | Not Specified
~~~

Report the demographics with the three columns separated by a pipe ‘|’ as a bar-separated row as above.

#### B.3 LLM prompt to extract list of diagnoses from PMOA case reports

##### Prompt

You are an expert physician. Output the list of diagnoses that pertain to the patient in the case report. For example, here is the case report.

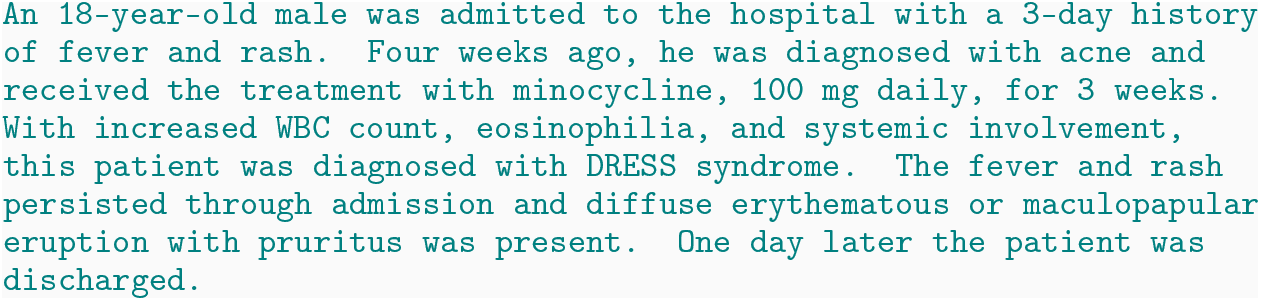

Then the output should look like:

~~~
acne
DRESS syndrome
~~~

Rash, leukocytosis, and other findings are not diseases so they are omitted, while acne and DRESS syndrome were both diagnosed, so they are included. Output a list of diagnoses (one per line) from the following case. Place the primary diagnosis first. Include only the list and nothing else.

### C Comparing LLM Annotations

In this section, we showcase an excerpt from PMC10629858, the latest case included in the sepsis-10 dataset [52]. To demonstrate the extraction task and analyze the performance of various LLM annotators, we compare manual annotations against outputs from several LLM variants: DeepSeek-

R1, DeepSeek-R1-UD-IQ1, DeepSeek-V3-0324, LLaMA 3.3 70B, OpenAI O1, OpenAI O3, and OpenAI O4-mini

Excerpt from [52]:

A 57-year-old man recently diagnosed with lepromatous leprosy was confirmed with skin biopsy and had been on treatment (rifampicin/clofazimine/dapsone) for 2 months before admission; he was presented to the hospital with complaints of abdominal distension, constipation, vomiting, and a 10-kg weight loss. On examination, the patient was vitally stable. He had evidence of peripheral lymphadenopathy with a distended abdomen and a positive shifting dullness. A computed tomography scan of his abdomen showed mural thickening of the terminal ileum with significantly enlarged mesenteric lymph nodes, mesenteric fat stranding, and intra-abdominal free fluid, suggesting abdominal granulomatous infection or neoplastic process….

The patient was planned for consolidation by autologous bone marrow transplant. Unfortunately, with the recurrent bacteremia and sepsis that accompanied the patient’s course due to his immunocompromised state, he was re-admitted to the medical ICU for severe sepsis and multiorgan failure and passed away around 6 months after his initial diagnosis with NHL, despite maintaining a remission status.

| Manual annotation |  | DeepSeek-R1 annotation |  |
| --- | --- | --- | --- |
| 57-year-old | 0 | 57 years old | 0 |
| man | 0 | male | 0 |
| diagnosed with lepromatous leprosy | -1461 | lepromatous leprosy | -1464 |
| on treatment | -1461 | skin biopsy | -1464 |
| rifampicin | -1461 | rifampicin | -1464 |
| clofazimine | -1461 | clofazimine | -1464 |
| dapsone | -1461 | dapsone | -1464 |
| presented to the hospital | 0 | admitted to the hospital | 0 |
| abdominal distension | 0 | abdominal distension | 0 |
| constipation | 0 | constipation | 0 |
| vomiting | 0 | vomiting | 0 |
| 10-kg weight loss | 0 | 10-kg weight loss | 0 |
| vitally stable | 0 | vital stability | 0 |
| peripheral lymphadenopathy | 0 | peripheral lymphadenopathy | 0 |
| distended abdomen | 0 | distended abdomen | 0 |
| positive shifting dullness | 0 | positive shifting dullness | 0 |
| computed tomography scan of his abdomen | 0 | computed tomography scan | 0 |
| mural thickening of the terminal ileum | 0 | mural thickening of the terminal ileum | 0 |
| significantly enlarged mesenteric lymph nodes | 0 | enlarged mesenteric lymph nodes | 0 |
| mesenteric fat stranding | 0 | mesenteric fat stranding | 0 |
| intra-abdominal free fluid | 0 | intra-abdominal free fluid | 0 |
| re-admitted to the medical ICU | 4383 | abdominal paracentesis | 0 |
| severe sepsis | 4383 | plan for autologous bone marrow transplant | 4320 |
| multiorgan failure | 4383 | recurrent bacteremia | 4320 |
| passed away | 4383 | sepsis | 4320 |
|  |  | readmission to ICU | 4320 |
|  |  | severe sepsis | 4320 |
|  |  | multiorgan failure | 4320 |
|  |  | death | 4320 |

### D Log-Time Cumulative Distribution Function

Recall the log-time cumulative distribution function is given as follows:

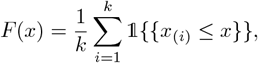

where 𝟙 is the indicator function.

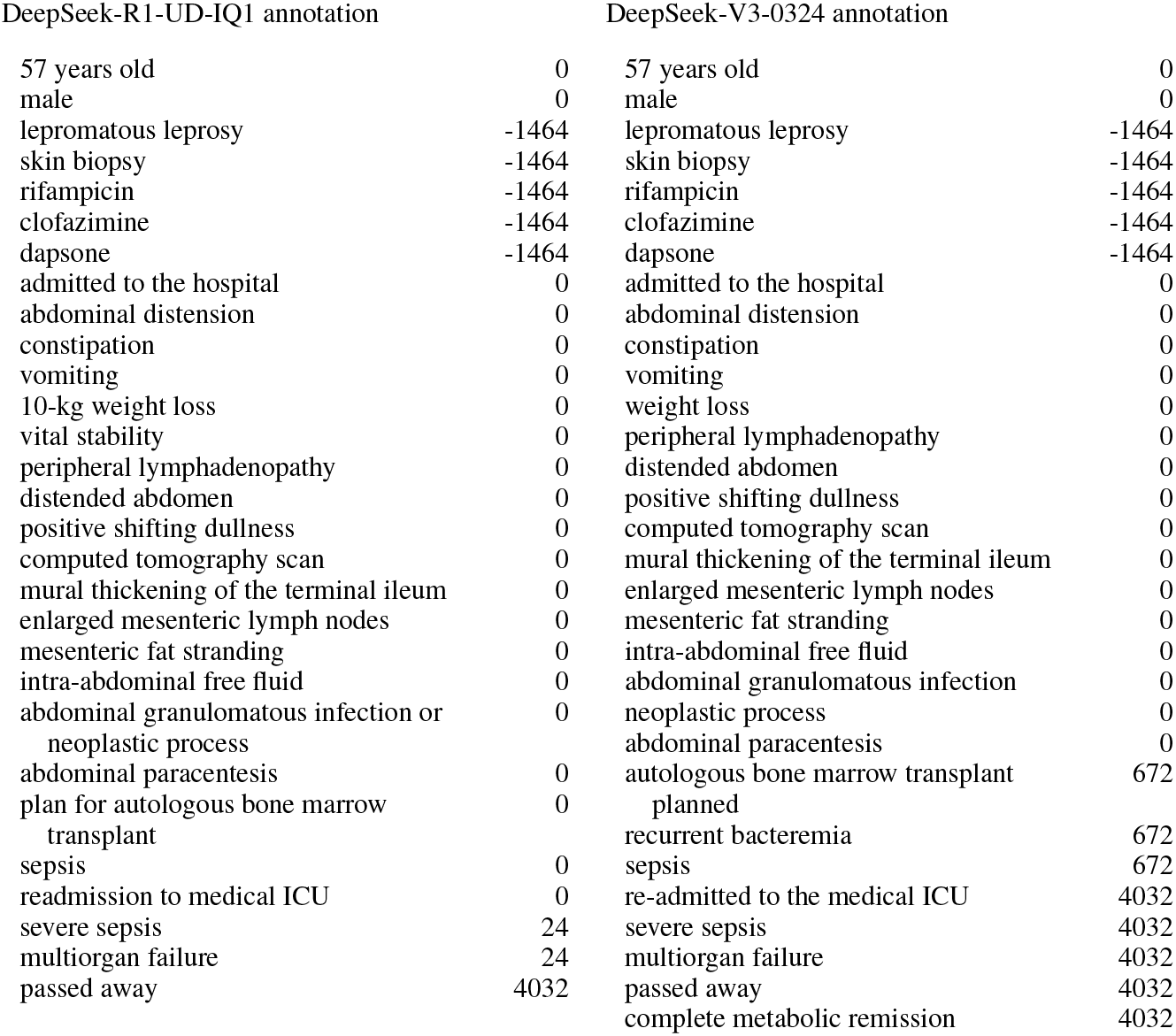

We compute the AULTC as the area under *F* (*x*) from *x* = 0 to *x* = log(1 + *S*_max_), normalized by log(1 + *S*_max_):

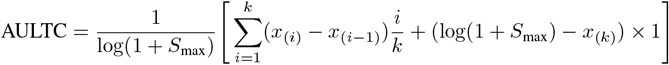

where *x*_(0)_ = 0. With this definition, AULTC = 1 indicates that discrepancies are zero (perfect recovery), resulting in maximum area log(1 + *S*_max_), and AULTC = 0 indicates that all discrepancies exceed *S*_max_, yielding zero area.

**Remark 1** *The time unit and cutoff S*_*max*_ *affect the AULTC calculation, so they must be specified when reporting the AULTC*.

Because of the log(1 +·) transformation (rather than the log(·) transformation which is undefined for zero time-error discrepancy), the discrepancies are non-linearly shifted in the log scale. The non-linear shift adjusts the relative widths of the step function (particularly for small discrepancies) changing the area calculation. The cutoff *S*_max_ affects the normalization factor. Therefore, these values should be chosen based on practicalities for the application. In our case, we chose hours at the time unit because the only sub-hour descriptions reported were several Apgar scores at the minute-level.

**Remark 2** *The average log-time discrepancy is non-convex*.

This can be seen from the observation that the log(1 +·) function is “spikier” than the L1 function. More formally, the log-time discrepancy has a higher curvature around zero error compared to the L1 loss. Consider the second derivatives of the loss functions with respect to the discrepancies

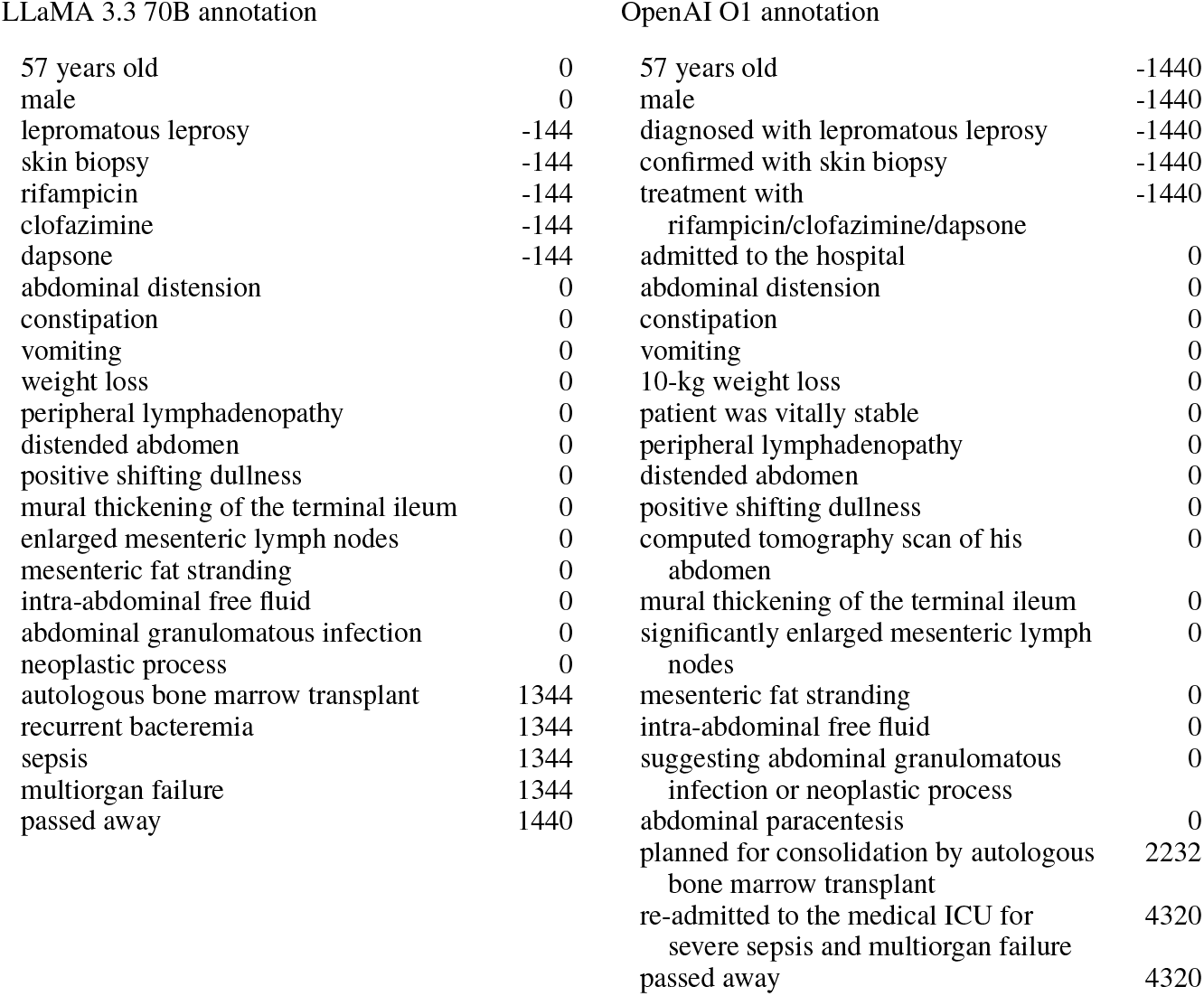

*s* = |*t*^*p*^ − *t*^*r*^|. For L1 loss *L* (*s*) = *s*, the second derivative 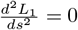 for *s* > 0. For the log-time loss, *L* (*s*) = log(1 + *s*), the second derivative is 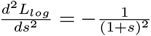. At *s* = 0, the second derivative is −1, while the second derivative of the L1 loss is 0.

To provide a simple counterexample, define the average log-time discrepancy is given by:

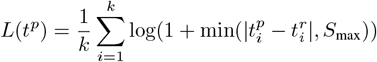

To prove that this function is non-convex, we need to show that there exist 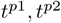 and *λ* ∈ (0, 1) such that

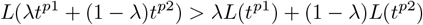

*L*(*λt*^*p*1^ + (1 − *λ*)*t*^*p*2^) > *λL*(*t*^*p*1^) + (1 − *λ*)*L*(*t*^*p*2^)

| − *× ×*

Consider the case where *k* = 1, *t*^*r*^ = 0, and *S*_max_ = 2. The loss function is *L*(*t*^*p*^) = log(1 + min(|*t*^*p*^|, 2)). Let 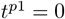 and 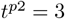. Choose *λ* = 0.5. Then 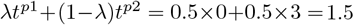. Now we evaluate the loss function at these points:

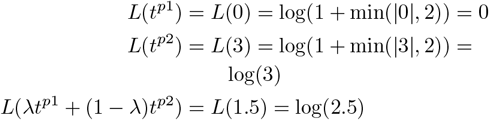

Now we check the convexity condition:

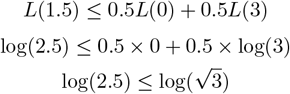

Since 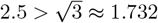, we have 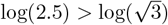. Therefore,

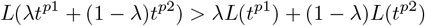

This proves that the average log-time discrepancy is non-convex. This remark illustrates the loss form, in case it is considered for optimization/model training rather than for assessment purposes.

**Figure C.1:**
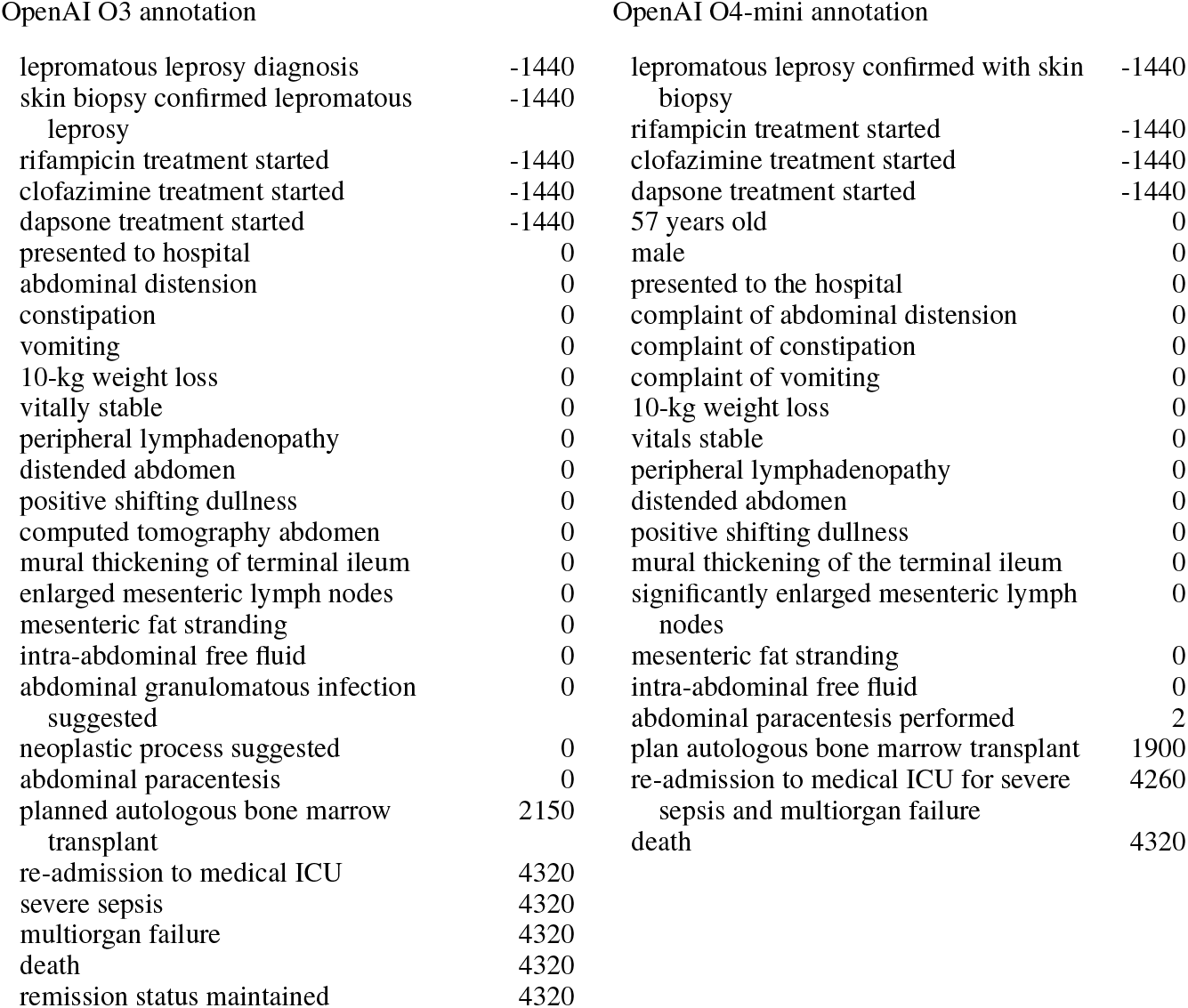
TTS comparison across multiple LLMs

### E Recursive Best Match Procedure

We provide pseudocode for the best match procedure between two lists of strings (Algorithm 1). For each list, we use the text order, *i*.*e*., the order of the events in the annotation files, to break embedding distance ties. We use the cosine similarity for the distance calculation using sentence transformer embeddings from `S-PubMedBert-MS-MARCO`.

### F Varying the Event Match Distance Threshold

By varying the event match distance threshold, we obtain different performance characteristics for event match rate, event concordance, and temporal discrepancy (AULTC). We choose a cosine similarity of 0.1 based on manual review of Llama 3.3 70B Instruct and o1 annotations. Plotting the event match rate versus temporal performance characteristic (concordance and AULTC) when varying the threshold from 0.01 to 0.25, we obtain performance tradeoffs in Figure F.1.

#### Algorithm 1

Recursive Best Match

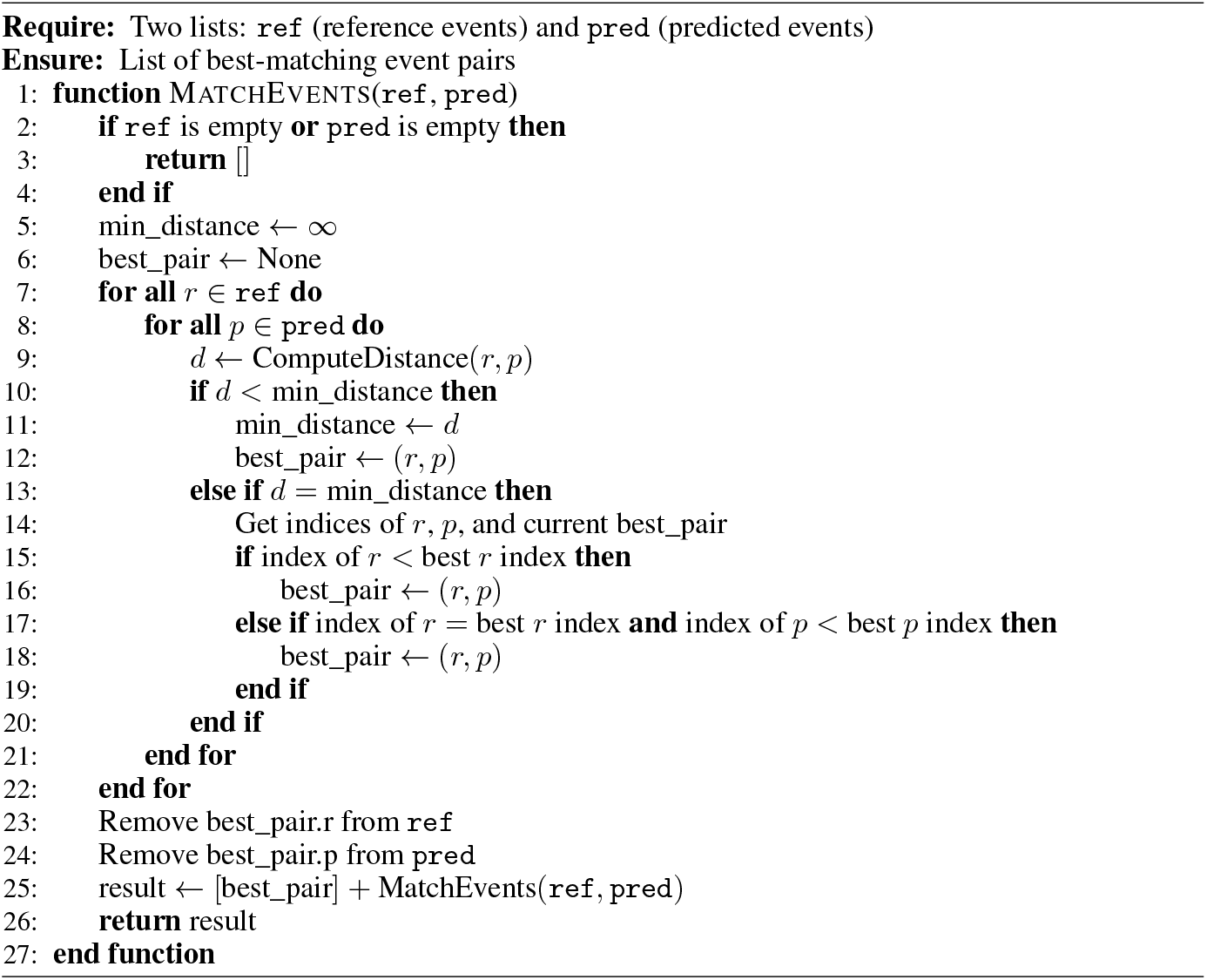

### G UMLS Mapping for Diagnosis Standardization

To standardize the diagnoses extracted by LLMs from the case reports, we mapped the free-text diagnoses to concepts in the Unified Medical Language System (UMLS). This process involved the following steps:

1. **Initial Extraction**: For each case report, we prompted the LLM to extract diagnoses as described in Appendix B.3. This yielded a list of free-text diagnoses for each case report.
2. **Entity Linking**: We used ScispaCy’s entity linker model `(en_core_sci_lg)` with the UMLS knowledge base to map each extracted diagnosis to corresponding UMLS Concept Unique Identifiers (CUIs). The entity linker identifies potential UMLS concepts and ranks them based on contextual relevance.
3. **Filtering and Disambiguation**: For each diagnosis, we:
  - Retained only mappings with a confidence score above 0.85
  - When multiple CUIs were identified for a single diagnosis, selected the highest-ranked mapping
4. **Concept Normalization**: After mapping to CUIs, we:
5. Retrieved the canonical names from UMLS Metathesaurus

This mapping process allowed us to standardize the diverse expressions of diagnoses extracted by LLMs into a consistent terminology system, enabling more reliable frequency analysis and co-occurrence patterns.

**Figure F.1:**
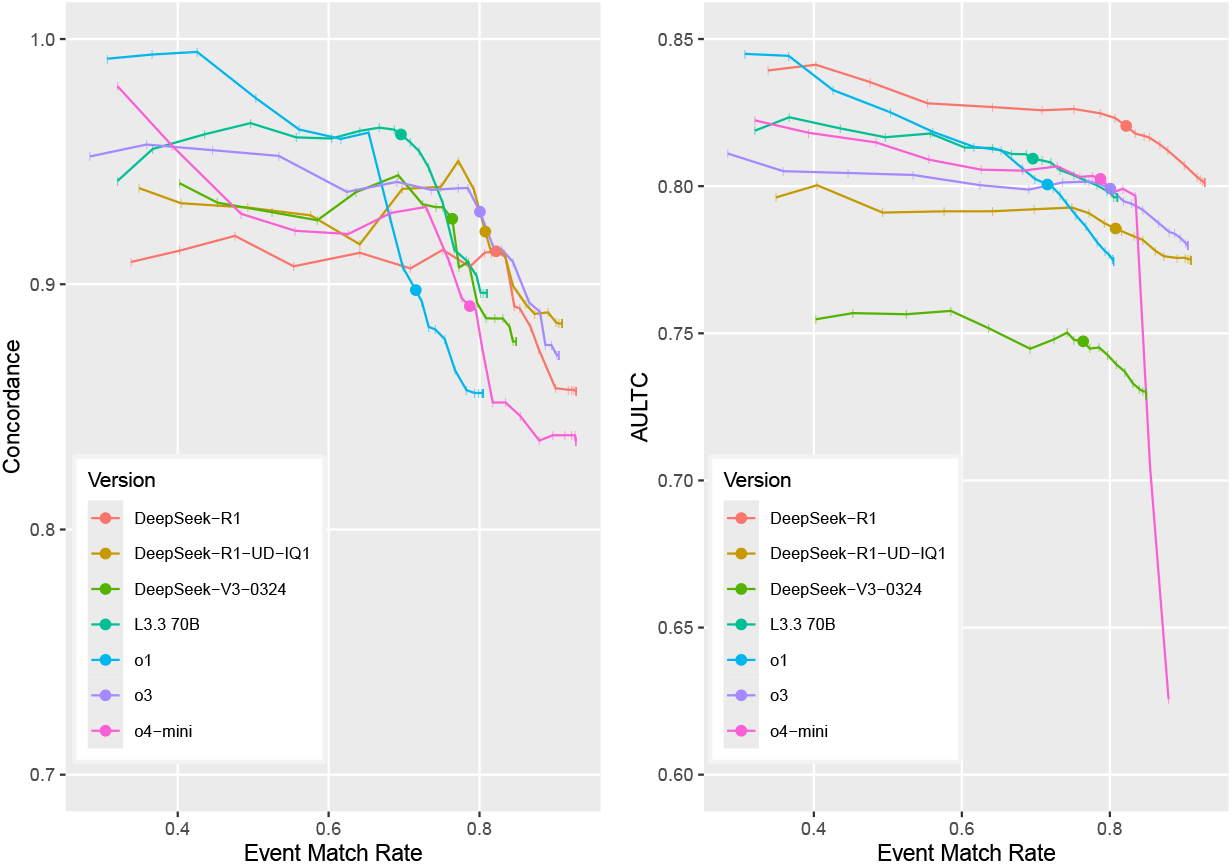
**Left:** Concordance by event match rate. Solid circle (*•*) represents threshold of 0.1, with ticks (|) indicating 0.01 increments of the threshold in [0.01, 0.25]. **Right:** AULTC by event match rate.

### H Survival Analysis Task on PMOA-TTS

We developed a survival analysis framework to evaluate the prognostic value of textual information encoded in the time series for time until death prediction. Our approach follows a two-stage process:

1. **Embedding Extraction**: Each textual time series is converted to a textual context in the format `“(time) clinical event [SEP] … [SEP] (time) clinical event [SEP]”` and processed using various pre-trained language models to obtain dense vector representations. For encoder models (BERT, RoBERTa, ModernBERT, etc.), we extract the `[CLS]` token embedding, while for decoder models (LLaMA, DeepSeek), we compute the mean-pooled embedding over all non-padding tokens.
2. **Survival Modeling**: These embeddings serve as covariates for two survival models:
  - **DeepSurv** [22]: A neural network generalization of the Cox proportional hazards model.
  - **DeepHit** [23]: A multi-task neural network jointly modeling discrete hazard and survival functions.

We evaluated the performance of the survival models using two metrics: the time-dependent concordance index [24], which measures the model’s ability to correctly rank survival times, and the integrated Brier score (IBS), which captures overall calibration and discrimination over time.

To ensure reliable estimates, we conducted experiments across five random seeds. For each seed, we performed a train-validation-test split with proportions of 64%, 16%, and 20%, respectively.

All survival times were right-censored at a fixed cutoff (`max_right_censoring_time`=1 year) to maintain consistency across samples. Only samples with positive survival times after censoring were included for model training and evaluation.

We tuned hyperparameters independently for each combination of language model and survival model. For DeepSurv and DeepHit, we searched over the following hyperparameter grid:

- `num_nodes`: {64, 128, 256, 512, 1024, 2048, 4096}
- `dropout`: {0.1, 0.5}
- `epochs`: 2000 (with early stopping)

For each configuration, the model was trained on the training set and evaluated on the validation set using the time-dependent concordance index. The configuration yielding the highest validation time-dependent concordance was selected, and the corresponding model was then evaluated on the held-out test set. This process was repeated for each random seed. We report the mean and standard deviation of the concordance-index and IBS across the five seeds for each combination of language model and survival model.

**Table H.1:**
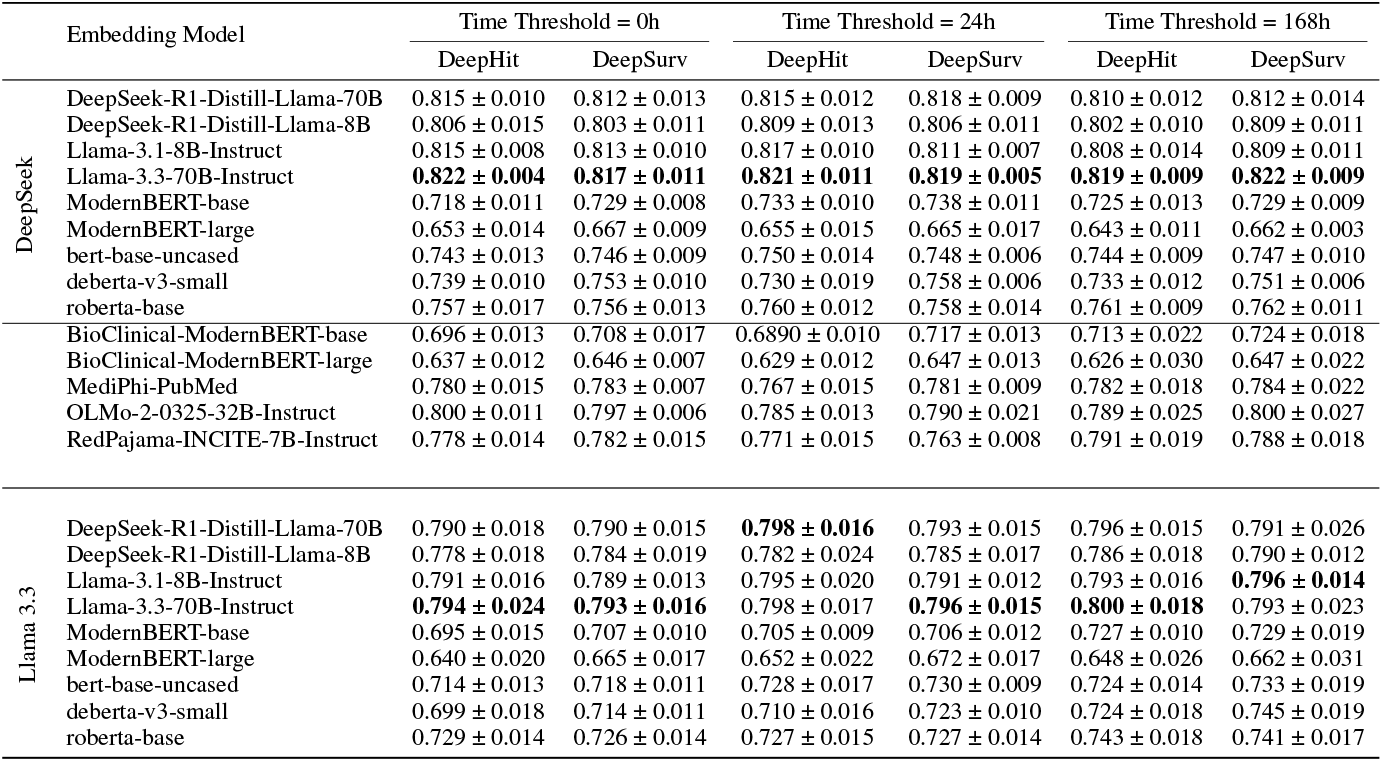
Time-dependent concordance (mean ± standard deviation across 5 experimental runs) index across embedding models and survival modeling approaches at different time thresholds for DeepSeek and Llama 3.3 annotations.

|  | Embedding Model | Time Threshold = 0h |  | Time Threshold = 24h |  | Time Threshold = 168h |  |
| --- | --- | --- | --- | --- | --- | --- | --- |
|  |  | DeepHit | DeepSurv | DeepHit | DeepSurv | DeepHit | DeepSurv |
| DeepSeek | DeepSeek-R1-Distill-Llama-70B | 0.815 ± 0.010 | 0.812 ± 0.013 | 0.815 ± 0.012 | 0.818 ± 0.009 | 0.810 ± 0.012 | 0.812 ± 0.014 |
| | DeepSeek-R1-Distill-Llama-8B | 0.806 ± 0.015 | 0.803 ± 0.011 | 0.809 ± 0.013 | 0.806 ± 0.011 | 0.802 ± 0.010 | 0.809 ± 0.011 |
| | Llama-3.1-8B-Instruct | 0.815 ± 0.008 | 0.813 ± 0.010 | 0.817 ± 0.010 | 0.811 ± 0.007 | 0.808 ± 0.014 | 0.809 ± 0.011 |
|  | Llama-3.3-70B-Instruct | **0.822 ± 0.004** | **0.817 ± 0.011** | **0.821 ± 0.011** | **0.819 ± 0.005** | **0.819 ± 0.009** | **0.822 ± 0.009** |
| | ModernBERT-base | 0.718 ± 0.011 | 0.729 ± 0.008 | 0.733 ± 0.010 | 0.738 ± 0.011 | 0.725 ± 0.013 | 0.729 ± 0.009 |
| | ModernBERT-large | 0.653 ± 0.014 | 0.667 ± 0.009 | 0.655 ± 0.015 | 0.665 ± 0.017 | 0.643 ± 0.011 | 0.662 ± 0.003 |
| | bert-base-uncased | 0.743 ± 0.013 | 0.746 ± 0.009 | 0.750 ± 0.014 | 0.748 ± 0.006 | 0.744 ± 0.009 | 0.747 ± 0.010 |
| | deberta-v3-small | 0.739 ± 0.010 | 0.753 ± 0.010 | 0.730 ± 0.019 | 0.758 ± 0.006 | 0.733 ± 0.012 | 0.751 ± 0.006 |
| | roberta-base | 0.757 ± 0.017 | 0.756 ± 0.013 | 0.760 ± 0.012 | 0.758 ± 0.014 | 0.761 ± 0.009 | 0.762 ± 0.011 |
| | BioClinical-ModernBERT-base | 0.696 ± 0.013 | 0.708 ± 0.017 | 0.6890 ± 0.010 | 0.717 ± 0.013 | 0.713 ± 0.022 | 0.724 ± 0.018 |
| | BioClinical-ModernBERT-large | 0.637 ± 0.012 | 0.646 ± 0.007 | 0.629 ± 0.012 | 0.647 ± 0.013 | 0.626 ± 0.030 | 0.647 ± 0.022 |
| | MediPhi-PubMed | 0.780 ± 0.015 | 0.783 ± 0.007 | 0.767 ± 0.015 | 0.781 ± 0.009 | 0.782 ± 0.018 | 0.784 ± 0.022 |
| | OLMo-2-0325-32B-Instruct | 0.800 ± 0.011 | 0.797 ± 0.006 | 0.785 ± 0.013 | 0.790 ± 0.021 | 0.789 ± 0.025 | 0.800 ± 0.027 |
| | RedPajama-INCITE-7B-Instruct | 0.778 ± 0.014 | 0.782 ± 0.015 | 0.771 ± 0.015 | 0.763 ± 0.008 | 0.791 ± 0.019 | 0.788 ± 0.018 |
| Llama 3.3 | DeepSeek-R1-Distill-Llama-70B | 0.790 ± 0.018 | 0.790 ± 0.015 | **0.798 ± 0.016** | 0.793 ± 0.015 | 0.796 ± 0.015 | 0.791 ± 0.026 |
| | DeepSeek-R1-Distill-Llama-8B | 0.778 ± 0.018 | 0.784 ± 0.019 | 0.782 ± 0.024 | 0.785 ± 0.017 | 0.786 ± 0.018 | 0.790 ± 0.012 |
| | Llama-3.1-8B-Instruct | 0.791 ± 0.016 | 0.789 ± 0.013 | 0.795 ± 0.020 | 0.791 ± 0.012 | 0.793 ± 0.016 | **0.796 ± 0.014** |
| | Llama-3.3-70B-Instruct | **0.794 ± 0.024** | **0.793 ± 0.016** | 0.798 ± 0.017 | **0.796 ± 0.015** | **0.800 ± 0.018** | 0.793 ± 0.023 |
| | ModernBERT-base | 0.695 ± 0.015 | 0.707 ± 0.010 | 0.705 ± 0.009 | 0.706 ± 0.012 | 0.727 ± 0.010 | 0.729 ± 0.019 |
| | ModernBERT-large | 0.640 ± 0.020 | 0.665 ± 0.017 | 0.652 ± 0.022 | 0.672 ± 0.017 | 0.648 ± 0.026 | 0.662 ± 0.031 |
| | bert-base-uncased | 0.714 ± 0.013 | 0.718 ± 0.011 | 0.728 ± 0.017 | 0.730 ± 0.009 | 0.724 ± 0.014 | 0.733 ± 0.019 |
| | deberta-v3-small | 0.699 ± 0.018 | 0.714 ± 0.011 | 0.710 ± 0.016 | 0.723 ± 0.010 | 0.724 ± 0.018 | 0.745 ± 0.019 |
| | roberta-base | 0.729 ± 0.014 | 0.726 ± 0.014 | 0.727 ± 0.015 | 0.727 ± 0.014 | 0.743 ± 0.018 | 0.741 ± 0.017 |

#### H.1 Survival Analysis Results for Different Annotations

We report results from both DeepSeek and Llama 3.3 (L33) annotations in Table H.1. Note that these annotation sources differ in construction and granularity. As such, the absolute concordance values are not directly comparable across annotation types. However, a consistent trend is observed across both: embeddings obtainted from larger LLMs, particularly Llama-3.3-70B-Instruct and DeepSeek-70B, generally achieve higher concordance scores, followed by their smaller variants and traditional transformer models like BERT and RoBERTa. This suggests that higher-capacity models may better encode the temporal progression patterns captured in textual time series, regardless of the specific annotation strategy used.

The relatively high time-dependent concordance values across both annotation sources indicate that textual representations of clinical event trajectories contain meaningful signal for survival prediction. This finding supports the viability of text-to-time modeling as a research direction. Future work can explore improved ways to encode textual time series–beyond simple embedding extraction and the triplet-based approach from [21]–with the goal of enhancing survival analysis performance and better understanding which modeling strategies are most effective for TTS. Additionally, these representations could eventually support alignment with structured data or serve as input to multimodal models in downstream clinical tasks. More broadly, these results suggest that clinical language models may serve as a foundation for temporal reasoning beyond survival analysis.

### I Forecasting Task on the Sepsis Subset of PMOA-TTS

To illustrate the potential of PMOA–TTS for downstream forecasting applications, we include previously reported results from a small subset of sepsis case reports [20]. In that study, LLM-extracted clinical textual time series (generated by Llama-3.3-70B) were used to train forecasting models for outcome prediction. While we do not rerun the experiment here, the relevant sepsis subset is now part of the publicly released PMOA–TTS corpus. This inclusion enables reproducibility of earlier findings and highlights how similar forecasting tasks can now be scaled across the full dataset or to other disease-specific cohorts. We define the following two main forecasting tasks.

#### Event Occurrence Prediction

In this task, the model receives a prefix of the clinical time-line—comprising all events up to a given time point *t*—and must predict whether each of the next *k* events will occur within a defined time window. This simulates an “online” prediction scenario, where, for each of the next *k* events, the model outputs a binary label: does this event happen within *h* hours after *t*? Time horizons considered include 1 hour, 24 hours (1 day), and 168 hours (1 week). The task is structured as a sequence of binary classification problems, with evaluation based on precision, recall, and F1 score, averaged across the *k* events for each time window.

#### Temporal Ordering Prediction

This task evaluates the model’s ability to infer the correct chronological sequence of future events. At each cutoff time *t*, we select the subsequent *k* events and remove their timestamps. The model is then required to output a permutation of these events that aligns with their true temporal order. This is formulated as a ranking problem, and performance is measured by the pairwise concordance—i.e., the proportion of correctly ordered event pairs—between the predicted and actual sequences. This setup challenges the model to infer temporal flow based solely on event content.

#### I.1 Modeling approaches

To evaluate performance on the event forecasting tasks, we use three modeling paradigms: (i) fine-tuned large language models (LLMs) with task-specific heads, (ii) prompted LLMs in zero- or few-shot settings without gradient updates, and (iii) BERT-style encoders with fine-tuned heads tailored to each task.

##### Fine-tuned LLMs

We use instruction-tuned decoder-only models from the `LLaMA` and `DeepSeek` families, including `Llama-3.3-70B-Instruct`, `Llama-3.1-8B-Instruct`, and their distilled variants. Each is paired with a lightweight MLP head for classification or ranking, depending on the task. Inputs include a text-formatted event prefix (e.g., timeline up to time *t*), optionally with an instruction template. The output layer generates task-specific predictions—binary labels for event occurrence or a permutation over *k* events for ordering—trained using cross-entropy or pairwise ranking loss.

##### Prompted LLMs

In the zero- or few-shot setting, we use the same LLM architectures without fine-tuning, guiding outputs via structured prompts at inference. Each prompt includes: (1) a system role (e.g., “You are an expert physician.”), (2) a task-specific user instruction, and (3) one or more few-shot examples. Prompts are tailored per task and described in [25]. Model outputs are parsed into binary labels or ordered lists; ambiguous or malformed generations are excluded from evaluation.

##### Fine-tuned Encoder-Only Models

We also evaluate encoder-based models trained end-to-end for each task, including `BERT-base-uncased`, `RoBERTa-base`, `DeBERTa-v3-small`, `ModernBERT-small`, and `ModernBERT-large`. Each is paired with a task-specific MLP head for event occurrence or ordering prediction. Inputs use the same tokenized event sequence prefixes as in other settings, formatted per model requirements. Models are trained with standard supervised objectives and evaluated using F1 score for event occurrence and pairwise concordance for ordering.

#### I.2 Forecasting results

We show the forecasting results for 2319 textual time series for sepsis (T2S2).[25]. From this set, we randomly selected two subsets for evaluation: a group of 10 reports (sepsis-10), which underwent expert clinical annotation to serve as a gold standard, and a larger group of 100 reports (sepsis-100) used for broader testing.

##### Event forecast within next 24 hours: F1 performance

Our results show that encoder-based models outperform LLMs in event forecasting across `Llama-3.3-70B` (Table **??**) annotations, especially in short- and medium-term prediction windows. Fine-tuned encoder models with MLP heads achieve significantly higher F1 scores than LLMs, confirming the strength of encoder-based representations for forecasting. Among them, MLP fine-tuning consistently outperforms masking-based methods, particularly for long-horizon forecasting. `ModernBERT-base` achieves the highest F1 across all horizons, especially at 168 hours, with `ModernBERT-large` performing similarly but without consistent gains. Zero-shot masking models like `BERT` and `RoBERTa` perform poorly, while `DeBERTa-small` and `ModernBERT-base` show stronger generalization.

**Table I.1:** Forecasting performance (event occurrence: F1 and correct event ordering: concordance-index) of the ensuing *k* = 8 events on the sepsis subset of PMOA-TTS (T2S2:**T**extual **T**ime-**S**eries for **S**epsis). **All models are trained/fine-tuned on time-ordered annotations from** `Llama-3.3-70B`. The sepsis-10 and sepsis-100 sets are evaluation subsets, consisting of 10 expert-annotated case reports serving as the gold standard and 100 sepsis reports for broader model evaluation respectively. Need to add new results for open-source and medically fine-tuned models.

|  | T2S2 (Llama 3.3) |  |  |  | sepsis-10 |  | sepsis-100 |  |
| --- | --- | --- | --- | --- | --- | --- | --- | --- |
|  | F1(1h) | F1(1d) | F1(1w) | c-index | F1(1d) | c-index | F1(1d) | c-index |
| **LLM-MLP head** |  |  |  |  |  |  |  |  |
| DS-L3.3 70B | 0.140 | 0.563 | 0.811 | **0.624** | **0.397** | **0.585** | **0.629** | **0.598** |
| L3.3 70B | 0.265 | 0.466 | 0.807 | 0.619 | 0.160 | 0.567 | 0.466 | 0.597 |
| DS-L3.1 8B | 0.264 | **0.584** | **0.837** | **0.624** | 0.258 | 0.576 | 0.605 | **0.598** |
| L3.1 8B | **0.338** | 0.565 | 0.823 | 0.619 | 0.368 | 0.569 | 0.536 | 0.596 |
| **Prompting** |  |  |  |  |  |  |  |  |
| L3.3 70B 0-shot | 0.085 | 0.443 | 0.616 | – | 0.236 | – | 0.452 | – |
| L3.3 70B 0-shot ordinal | 0.063 | 0.454 | 0.707 | – | 0.289 | – | **0.483** | – |
| L3.3 70B few-shot | **0.099** | 0.457 | 0.703 | – | **0.360** | – | 0.452 | – |
| L3.3 70B few-shot ordinal | 0.064 | **0.460** | **0.729** | – | 0.313 | – | 0.480 | – |
| **Encoder-MLP head** |  |  |  |  |  |  |  |  |
| BERT | 0.224 | 0.622 | 0.899 | 0.500 | 0.400 | 0.500 | 0.612 | 0.500 |
| RoBERTa | 0.180 | 0.608 | 0.900 | 0.500 | 0.364 | 0.500 | 0.597 | 0.500 |
| DeBERTa-small | 0.258 | 0.641 | 0.898 | **0.603** | 0.427 | 0.585 | 0.616 | **0.592** |
| ModernBERT-base | 0.257 | **0.645** | **0.903** | 0.594 | **0.431** | **0.590** | **0.626** | 0.565 |
| ModernBERT-large | **0.312** | 0.622 | 0.897 | 0.500 | 0.404 | 0.500 | 0.623 | 0.500 |
| **Encoder-masking-fine-tuned** |  |  |  |  |  |  |  |  |
| BERT | 0.131 | 0.471 | **0.741** | 0.622 | 0.320 | 0.580 | 0.467 | 0.622 |
| RoBERTa | 0.129 | 0.484 | 0.659 | 0.625 | 0.329 | 0.587 | 0.472 | 0.614 |
| DeBERTa-small | 0.135 | 0.468 | 0.693 | **0.648** | 0.318 | **0.602** | 0.462 | 0.623 |
| ModernBERT-base | **0.139** | **0.489** | 0.700 | 0.632 | **0.334** | 0.595 | **0.478** | 0.612 |
| ModernBERT-large | 0.124 | 0.470 | 0.712 | 0.646 | 0.318 | 0.586 | 0.468 | **0.631** |
| **Encoder-masking-zeroshot** |  |  |  |  |  |  |  |  |
| BERT | 0.000 | 0.000 | 0.000 | 0.504 | 0.000 | 0.515 | 0.000 | 0.506 |
| RoBERTa | 0.074 | 0.279 | 0.445 | 0.500 | 0.083 | 0.500 | 0.319 | 0.500 |
| DeBERTa-small | 0.008 | 0.439 | 0.839 | **0.562** | 0.296 | **0.518** | **0.518** | 0.501 |
| ModernBERT-base | **0.132** | **0.525** | **0.840** | 0.556 | **0.352** | 0.503 | 0.511 | 0.501 |
| ModernBERT-large | 0.109 | 0.413 | 0.622 | 0.499 | 0.326 | 0.509 | 0.339 | **0.518** |

Performance improves with longer forecasting windows, indicating better predictability of long-term trends and greater difficulty in short-term forecasting due to event sparsity. Models also generalize poorly to external datasets (sepsis-10, sepsis-100), especially at the 1-day horizon, highlighting challenges in domain adaptation. Nonetheless, `ModernBERT` maintains strong relative performance, demonstrating robustness across datasets.

##### Temporal ordering of forecasted events: Concordance

In addition to event occurrence prediction, models are evaluated on their ability to rank upcoming events using concordance (c-index), based on annotations from `Llama-3.3-70B` (Table **??**). Encoder models again outperform LLMs, which fail to produce reliable orderings even with fine-tuning or prompting. Within encoder architectures, training strategies impact performance: MLP-head fine-tuned models lead in F1 but do not consistently achieve the best concordance. Instead, fine-tuned masking models—particularly `ModernBERT-large`—achieve the highest c-index, indicating a strength in capturing event order over binary classification.

Dataset comparisons reveal generalization challenges. While fine-tuned models perform well on the internal T2S2 set, c-index scores drop on external datasets, especially for non-`ModernBERT` models. `ModernBERT-large` (fine-tuned masking) maintains top c-index scores across all datasets, confirming its robustness. MLP-head models retain strong F1 scores but slightly lag in concordance, especially at the 168-hour horizon, suggesting trade-offs between classification and ranking.

Overall, `ModernBERT` models are the strongest across F1 and concordance, with MLP-head finetuning excelling at binary tasks and masking fine-tuning better suited for event ordering. However, all models struggle with external generalization, underscoring the need for improved domain adaptation.

### J Computational Resources for Reproducibility

We designed the PMOA–TTS pipeline to ensure reproducibility and transparency across all stages of extraction, annotation, and evaluation. All experiments and annotations involving large language models (LLMs) were executed using commercially available APIs (OpenAI) or open-source models run on GPU-accelerated infrastructure (HuggingFace).

For LLM-based extraction and classification tasks (e.g., timeline creation, diagnosis extraction, and case report filtering), we primarily used DeepSeek-R1, LLaMA 3.3–70B Instruct, and OpenAI models (O1, O3, O4-mini). DeepSeek-R1 was run on an 6× NVIDIA H200 GPUs. Inference for DeepSeek-R1-IQ1 and LLaMA 3.3–70B were performed using Hugging Face’s transformers library on a 2×A100 GPU setup. Each inference pass over a full case report required between 10 to 120 seconds per document depending on model size, context length, and prompt complexity. Total timeline extraction across 124,699 reports took approximately 8400 GPU-hours on DeepSeek-R1 and 2000 GPU-hours for LLaMA 3.3–70B. Temperature was set to recommended levels of 0.6 for DeepSeek models, and 0.7 for Llama 3.3-70B. Experiments using OpenAI’s hosted models were conducted via the OpenAI API. The prompt templates and exact settings are available in Appendices B.1, B.2 and B.3.

For embedding generation in the survival modeling task (Section 4.5), decoder models (LLaMA, DeepSeek) used mean-pooled hidden states, and encoder models (e.g., PubMedBERT) used [CLS] embeddings. Survival models (DeepSurv, DeepHit) were trained using PyTorch on single NVIDIA GeForce RTX 4090. Each training run optimizing across the hyperparamater grid for the validation set took on average 195 minutes. Hyperparameter search across seeds, dropout values, and hidden node counts (Appendix H) resulted in 48 GPU-hours total compute.

Our full codebase, including preprocessing scripts, prompt templates, and evaluation pipelines, is available at: https://github.com/jcweiss2/pmoa_tts. The annotated dataset is hosted at: https://huggingface.co/datasets/snoroozi/pmoa-tts. Instructions for reproducing all main results, including model prompts and evaluation metrics, are included in the dataset’s documentation.

We emphasize that all models used were publicly available or accessible via API, and no proprietary infrastructure was required. This ensures that all reported results can be reproduced with modest compute resources by academic researchers with access to common cloud platforms or institutional GPUs.

### K Sensitivity to the Quality of LLM Prompts

For sensitivity analyses with respect to the quality of different prompt strategies for extracting textual time-series (DeepSeek R1) from PMOA case reports, we omitted selected sections of the LLM query prompt (see Appendix B.1 for the full prompt). This led to the following ablations.

1. **No ablation (DSR1)**: Original LLM query prompt for DeepSeek R1.
2. **No role:** Remove the sentence `“You are a physician.”`
3. **Zero-shot prompting:** Remove the example case report and textual time-series from the prompt. `An 18-year-old male was admitted to..` “Let’s find the locations of event in the case report, it shows that … then the output should look like:” `18 years old | 0` `male | 0` ..
4. **No conjunction expansion:** Remove the following sentence from the prompt: `“Separate conjunctive phrases into its component events and assign them the same timestamp (for example, separation of ‘fever and rash’ into 2 events: ‘fever’ and ‘rash’)”`
5. **Interval:** A query was added to the prompt for extracting the time interval (start and end time of clinical events. The original prompt’s instructions and few-shot examples were modified accordingly.
6. **Interval + Type:**. A query was added to the prompt for extracting the ime interval and i2b2 event type (Interval+Type), where event type is one of: Factual, Possible, Hypothetical, Conditional, Negated, Historical, Uncertain. The original prompt’s instructions and few-shot examples were modified accordingly.

**Figure K.1:**
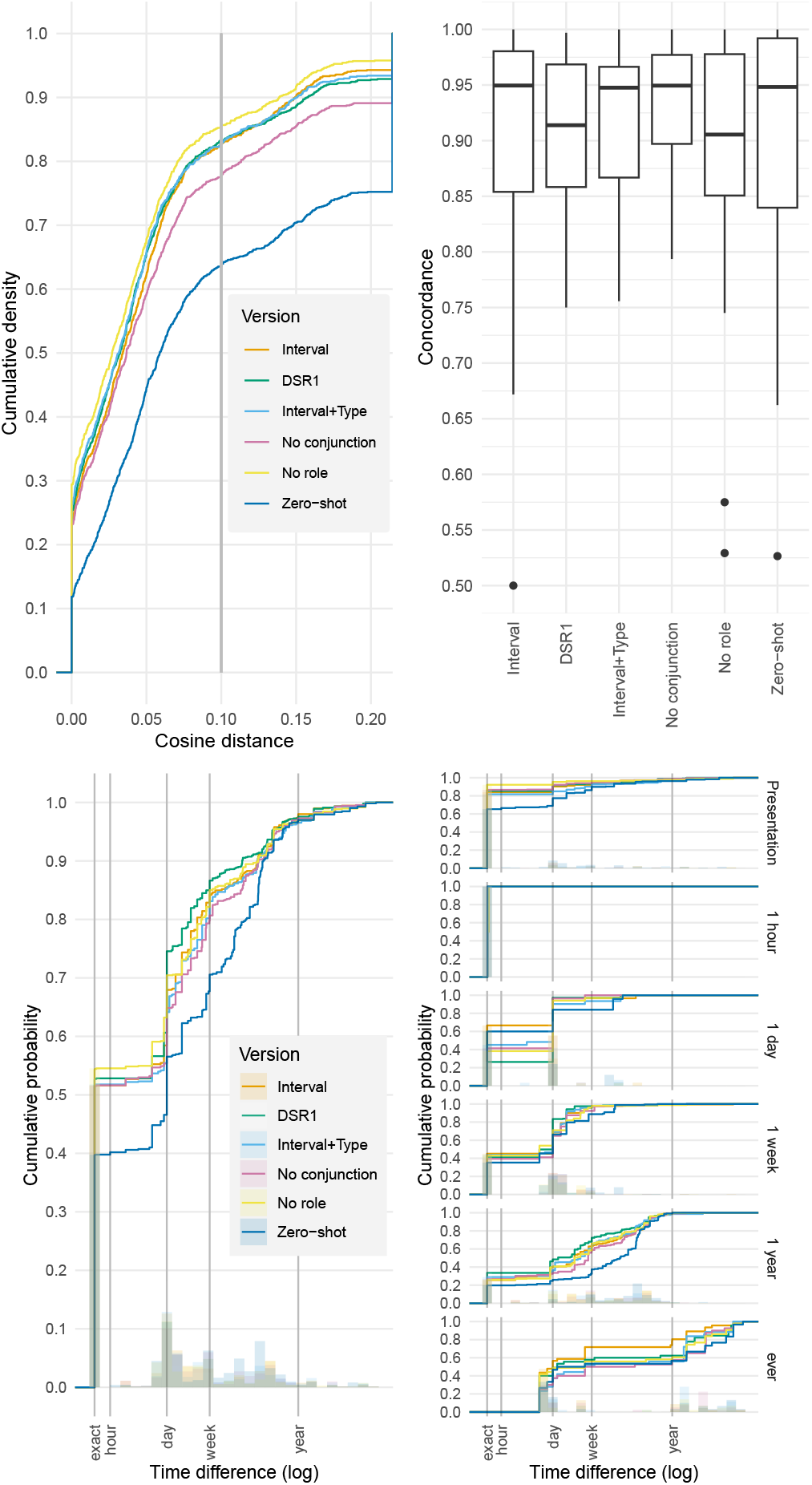
Event match cumulative distribution function (left), concordance box plots (middle left), time discrepancy from the manual annotation timestamps among matched events, overall (middle right), and disaggregated by clinician annotator timestamp (time from presentation, right). Each curve represents a different prompt ablation or variant used for extracting temporal clinical events from case reports.

Figure K.1 presents sensitivity analyses for different prompt strategies in extracting structured time-series from clinical narratives. Each ablation tests the effect of removing specific instructional components or adding structured extraction requirements. We evaluate event-level semantic similarity, timestamp accuracy, and temporal concordance across matched events.

#### Semantic similarity

The cumulative density of cosine distances between matched events reveals that omitting key components of the prompt negatively impacts text alignment (Figure Figure K.1, far left). The Interval, DSR1, and Interval+Type prompts consistently show high-quality event matches, with over 80% of events falling below a cosine distance of 0.1. In contrast, removing role framing (No role) or conjunction splitting (No conjunction) leads to modest degradation. The Zero-shot condition—lacking both examples and guiding instructions—performs markedly worse, with only 60% of events under the same similarity threshold.

#### Temporal concordance

Concordance index (c-index) measures the agreement between predicted and annotated event ordering (Figure Figure K.1, middle left). The Interval+Type variant achieves the highest median concordance, followed closely by Interval, DSR1, and No role. Removing conjunction expansion reduces ordering consistency slightly, while Zero-shot again yields the lowest performance with a wider spread and multiple outliers (c-index < 0.6), indicating inconsistent temporal sequencing.

#### Timestamp accuracy

Cumulative distributions of time discrepancies from manual annotations further support these findings (Figure K.1, middle right). Structured prompts—especially Interval, DSR1, and Interval+Type—demonstrate tighter timestamp alignment. Ablations involving role removal or conjunction omission produce broader error distributions, and Zero-shot extraction results in substantially larger timestamp mismatches.

When stratified by the time from presentation (e.g., within 1 hour, 1 day, 1 week), prompt differences are most pronounced for events occurring farther from the report anchor (Figure K.1, far right). For immediate events, all prompt versions perform comparably. However, at longer timescales (1 week to “ever”), Interval and Interval+Type maintain superior accuracy, while Zero-shot shows dramatic drops in performance. This suggests that structured prompts are particularly critical for accurate long-range temporal reasoning.

### L Broader Impact

The PMOA–TTS corpus contributes to the broader goals of equitable, scalable, and transparent medical AI. However, it also introduces societal considerations related to the use of LLM-generated clinical timelines, especially in sensitive healthcare contexts. Below, we discuss the potential positive and negative societal impacts of our work.

The PMOA–TTS corpus offers several potential positive contributions to the biomedical NLP and clinical AI communities. By releasing the largest openly available dataset of temporally structured clinical narratives, this work democratizes access to timeline-based clinical text—an area typically limited to proprietary EHR datasets. The corpus spans a wide range of diagnoses and demographics, supporting research on temporal patterns in diverse and sometimes underrepresented conditions. Additionally, by making the extraction pipeline, prompting strategies, and evaluation framework publicly available, the project promotes transparency and reproducibility in LLM-based clinical information extraction. These contributions collectively advance the development of models for timeline reconstruction, temporal reasoning, and longitudinal prediction, while lowering the barrier to entry for researchers working in resource-limited settings.

At the same time, the release and use of PMOA–TTS raise important considerations. Although LLMs achieve strong performance, they may still generate inaccurate or hallucinated outputs, and any downstream use of these annotations without further validation could introduce bias or error.

The extraction of structured diagnoses, demographics, and clinical events from public text—though non-identifiable—could also lead to misinterpretation or reinforce historical biases if not handled carefully. Furthermore, while the dataset is intended for research, there is a risk of misapplication in high-stakes clinical settings without expert oversight. Lastly, the stylistic and structural differences between case reports and real-world EHR notes may limit the generalizability of findings derived from PMOA–TTS. These limitations underscore the importance of using the corpus responsibly, with clear reporting of model limitations and appropriate safeguards when applied in downstream tasks.

